# Efficient Classification of Pulmonary Pneumonia and Tuberculosis Alongside Normal and Non-X-ray Images with Minimal Resources and Maximum Accuracy

**DOI:** 10.1101/2024.12.31.24319820

**Authors:** Rifatul Islam Majumder

## Abstract

Pneumonia, primarily caused by *Streptococcus pneumoniae*, and tuberculosis (TB), caused by *Mycobacterium tuberculosis*, continue to present significant global health challenges. Pneumonia is responsible for 14% of deaths among children under five, resulting in 740,180 fatalities annually [1]. Similarly, TB caused 1.25 million deaths in 2022, including 161,000 among individuals with HIV [2]. Misdiagnosis is a critical issue, with 22.3% of pneumonia cases being misidentified as TB [3], highlighting the need for accurate diagnostic tools. This study proposes a novel classification framework for chest X-ray (CXR) images, designed to identify four categories: normal, pneumonia, tuberculosis, and non-X-ray. By incorporating the “non-X-ray” class, the model enhances robustness by detecting outliers and unseen anomalies. The framework utilizes a pre-trained ResNet-18 convolutional neural network and a fine-tuned DenseNet-121, both trained with and without weighted loss function. The best-performing model achieved exceptional results, with 98.76% accuracy, 99.01% precision, and 99.03% recall, maintaining or surpassing class-wise performance. The model was trained on a curated dataset from multiple valid sources, containing 5,489 normal, 4,273 pneumonia, 4,197 tuberculosis, and 1,357 non-X-ray images. This framework has the potential to reduce misdiagnosis and improve healthcare delivery, particularly in resource-limited environments.

## 1.2) Introduction

Pneumonia and tuberculosis (TB) pose substantial global health challenges, particularly in low-resource settings where diagnostic tools are limited. Pneumonia, predominantly caused by *Streptococcus pneumoniae*, contributes to 14% of deaths in children under five, with 740,180 fatalities annually [1]. It also ranks as the second-leading cause of hospitalization among Medicare beneficiaries, with over 600,000 admissions yearly [4], [5]. TB, caused by *Mycobacterium tuberculosis*, accounted for 1.25 million deaths in 2022, including 161,000 among people living with HIV, with an estimated global incidence of 10.8 million cases [2].

Misdiagnosis remains a critical concern. Studies report that 22.3% of pneumonia cases are misdiagnosed as pulmonary TB [3], leading to treatment delays and increased mortality. Untreated pneumonia has a mortality rate of up to 20% [6], while untreated TB fatalities range from 4.8% to 60% depending on disease stage and co-morbidities [2], [7]. These diagnostic challenges highlight the need for automated systems capable of accurately distinguishing between these diseases.

The advent of artificial intelligence (AI) has enabled the development of machine learning (ML) and deep learning (DL) algorithms capable of addressing complex diagnostic challenges in medical imaging. Algorithms such as logistic regression, decision trees, and neural networks—modeled on biological neurons—have advanced to more sophisticated DL approaches, forming the basis of groundbreaking frameworks like ResNet, DenseNet, and Vision Transformers [8], [9]. These tools have proven highly effective for tasks such as lung segmentation, disease classification, and treatment prediction, often achieving results comparable to or exceeding human performance [10], [11]. CNNs allow data-driven, highly representative, hierarchical image features to be learned from adequate training data, but obtaining data sets in the medical imaging domain as comprehensively annotated, as ImageNet remains a challenge [12]–[14]. It is also worth noting that the healthcare sector is entirely different from any other field being a high priority sector with customers willing to pay for highest quality of care and services. The healthcare sector has not fulfilled the aspirations of society, while the industry absorbs a large percentage of national budgets. The medical experts examining medical data which substantially suffer from subjective differences and the quality of the images and by the fatigue caused due to heavy workload. Thus, application of machine learning in health care sector is gathering a lot of attention in recent times. Artificial intelligence (AI) based solutions have been proposed for many biomedical applications including brain tumor, lungs nodule, pneumonia, breast cancer detection, physiological monitoring and social sensing [15]–[20]. Among the deep machine learning (ML) techniques, convolutional neural networks (CNNs) have shown great promise in image classification and therefore widely adopted by the research community [9], [21]-[26]. X-ray radiography is a low-cost imaging technique and there is an abundance of data available for training different machine learning models making deep learning techniques popular for the automatic detection of lung diseases from chest radiographs. Moreover, we have a team of medical doctors who are experts in interpreting chest X-ray images put us in a favourable condition for computer-aided diagnostic system development.

CNNs have been used in several recent studies for the detection of lungs diseases including pneumonia and tuberculosis by analysing chest X-ray images. In response to the COVID-19 pandemic situation in 2020, CNN based techniques have been used for the detection of the novel coronavirus infection form CXR images. Tahir *et al.* [27] classified different coronavirus families (SARS, MERS and COVID-19) using transfer learning of various pre-trained CNN models with sensitivity values greater than 90%. Chowdhury *et al.* [28] developed a trained model using chest X-ray dataset to distinguish COVID-19 pneumonia, viral pneumonia and normal cases.

However, current models often neglect the identification of “non-X-ray” or anomalous cases, a critical requirement for real-world clinical settings [9], [21]-[25]. Moreover, no prior study has explored multi-class classification encompassing pneumonia, TB, and non-X-ray cases while accounting for outliers.

This study proposes a novel multi-class classification system using pretrained CNN model that categorizes CXRs into four classes: normal, pneumonia, tuberculosis, and non-X-ray. By incorporating the “non-X-ray” category, our model enhances reliability and robustness.

The rest of the paper is divided in the following sections: Section 2 summarizes current state-of-the-art works, limitation of the state-of-the-art works. Section 3 describes explanation of this approach and validity of this approach, dataset and custom dataset splits. Section 4 for methodology of this study, while Section 5 summarizes the model results, using Grad-CAM for visualization, Direct comparison with state-of-the-art works, Deployment strategy discussion. Finally, Section 6 research outcome, conclusion.

### 1.3) Ethics Statement

This study adhered to all relevant ethical guidelines. All data used in this study were anonymized and publicly available for research purposes. No additional ethical approval was required as the datasets utilized are open-access, and their use for research has been authorized by the original data sources. The study was conducted in accordance with the Declaration of Helsinki.

## Section 2: Related Work & Limitations

### 2.1) Current state-of-the-art works

Rahman et al. [25] developed a framework for reliable tuberculosis (TB) detection from chest X-rays by incorporating image preprocessing, data augmentation, UNet segmentation, and deep learning. Their model, tested on a dataset of 3,500 TB-infected and 3,500 normal X-rays, utilized nine pre-trained convolutional neural networks (CNNs), including ResNet-18, DenseNet-121, and DenseNet-201. After segmenting lung images, the model achieved 98.6% accuracy with DenseNet-201, accompanied by Grad-CAM visualization.

Sharma et al. [24] developed a deep learning-based TB detection model using the UNet segmentation and Xception classification models. The model achieved 96.35% accuracy for segmentation and 99.29% accuracy for classification. Using Grad-CAM, the model visualized lesions in the lungs.

Sharma and Guleria [21] proposed a VGG16-based model for pneumonia detection, tested on two datasets. Moreover, large portion of each dataset derives from Kermany dataset [29]. The model achieved 92.15% accuracy on the first dataset and 95.4% on the second, outperforming other classifiers like SVM and KNN. Their work highlighted the model’s robust performance in pneumonia classification.

Singh et al. [9] introduced a pneumonia detection framework using the Vision Transformer (ViT) on a Kaggle chest X-ray dataset. The ViT model, leveraging self-attention mechanisms, outperformed CNNs, achieving 97.61% accuracy, 95% sensitivity, and 98% specificity. Their work set a new performance benchmark in pneumonia detection.

Kundu et al. [22] proposed an ensemble deep learning model combining GoogLeNet, ResNet-18, and DenseNet-121 for automatic pneumonia detection. Using a novel weighted ensemble approach, the model achieved impressive results on two publicly available datasets. On the Kermany dataset [29], it obtained an accuracy of 98.81%, with a sensitivity of 98.80%, a precision of 98.82%, and an F1-score of 98.79%, demonstrating strong performance in classifying pneumonia cases. However, when evaluated on the RSNA dataset [30], the model’s accuracy dropped to 86.85%, with a sensitivity of 87.02%, precision of 86.89%, and F1-score of 86.95%.

Singh and Tripathi [23] proposed a quaternion-based deep learning model for pneumonia classification from chest X-rays. They used a Quaternion Convolutional Neural Network (QCNN), a generalization of conventional CNNs, which treats all three-color channels (R, G, B) as a single unit. This approach improves feature extraction and classification. Their model, trained on a large Kaggle dataset, achieved 93.75% accuracy and an F1-score of 0.94.

Kabir et al. [31] applied knowledge distillation techniques to enhance real-time detection of COVID-19, pneumonia, and tuberculosis from chest X-rays by transferring knowledge from a larger “Teacher” model to a smaller “Student” network. This approach reduced computational complexity, enabling deployment in mobile and cloud-based platforms while achieving high performance. Trained on a combined dataset of chest radiographs, the model achieved 97% accuracy, with 94% precision and 97% recall, specific results for the student model included precision/recall scores of 95%/97% for COVID-19, 94%/94% for pneumonia, 93%/98% for tuberculosis, and 98%/98% for normal cases.

Ahmed et al. [32] proposed a hybrid feature-based approach for differentiating pneumonia and tuberculosis using X-ray images. Their work addressed the critical challenge of distinguishing these diseases early due to their overlapping radiological features. They used a combination of VGG16, ResNet18, and feature extraction methods like Local Binary Patterns (LBP), Discrete Wavelet Transform (DWT), and Gray Level Co-occurrence Matrix (GLCM). Their most promising system, combining VGG16 features with LBP, DWT, and GLCM, achieved 99.6% accuracy, 99.17% sensitivity, and 99.63% precision. They also utilized Principal Component Analysis (PCA) to reduce feature dimensionality before classification with an artificial neural network (ANN).

### 2.2) Limitations of the current state-of-the-art

Kabir et al. [31] proposed a distillation technique to reduce the computational complexity of deep learning models by training smaller “student” models to replicate the performance of larger “teacher” models.

However, this approach has notable limitations, especially in medical applications. The student models in their study, with 10 million parameters, are comparable in depth to a densenet-121 model. In fact, this setup offers no significant computational advantage over directly training a ResNet-18 or similar models like DenseNet-121. Additionally, the TB dataset used in the study, curated by Tawsifur Rahman from Kaggle [33], theoretically integrates data from multiple sources, including RSNA [30], NIAID [34], Shenzhen [35], Montgomery [35], and Belarus [36]. However, as of December 22, 2024, the cross-validation was limited to data from only RSNA [30] and 20% of NIAID [34], which significantly reduces the model’s generalizability on tuberculosis classification and raises concerns about the validity of the results. While Kabir’s method is computationally efficient and it considers about Covid-19 detection too by using 3616 samples, it does not address important challenges such as domain shift, knowledge stability, and catastrophic forgetting. On the other hand, Zhu et al.’s [37] tri-enhanced distillation framework offers a more robust solution by incorporating stochastic knowledge augmentation, adaptive knowledge transfer, and global uncertainty-guided fusion, which better handle dynamic datasets and improve model stability and generalization, making it more suitable for medical applications than Kabir’s approach.

Ahmed et al. [32] demonstrated the effectiveness of hybrid techniques for distinguishing between pneumonia and tuberculosis (TB) using X-ray images. However, their study also suffers from similar limitations. Notably, it relies on the same imbalanced dataset curated by Tawsifur Rahman from Kaggle [33], where certain classes, such as tuberculosis, are underrepresented. While data augmentation methods were applied to address this imbalance, artificially increasing the dataset may not accurately reflect real-world medical distributions. Moreover, the use of CNNs like VGG16 and ResNet-18 in combination with handcrafted features (e.g., LBP, DWT, and GLCM) may limit the models’ robustness and generalizability across diverse datasets and populations. The small size of the medical datasets also raises concerns about overfitting. Additionally, the computational complexity of these hybrid systems may hinder their scalability and efficiency in real-world clinical settings, limiting their practical application. Since the study is based on a single dataset, the models are likely to perform poorly when applied to other healthcare environments or medical imaging sources, further restricting their generalizability.

The study by Rahman et al. [25] also has several limitations. The lack of ground truth masks for the segmentation database makes it difficult to rigorously evaluate the segmentation model, which could undermine confidence in its generalization to new, unseen data. Furthermore, the study relies on the same Kaggle dataset used by both Kabir et al. [31] and Ahmed et al. [32], raising concerns about data integrity and its ethical value. While the study reports high performance metrics, it does not address critical issues like false negatives and false positives in tuberculosis diagnosis. Finally, the computational resource demands of deep learning models such as DenseNet201 and CheXNet may limit their applicability in resource-constrained settings, such as rural or low-resource healthcare environments. The work is a binary classification and in paper authors are also concerned about the unknown class or non-X-ray images which is crucial for real world scenarios.

Sharma et al. [24] attempted to address data transparency concerns raised by Rahman et al. [25] but can arise similar UNet segmentation overfitting over current datasets and minimal utilization of the NIAID [34] dataset and a small sample size limit the generalizability of their findings. Additionally, the study’s focus on binary classification overlooks the critical need to address non-X-ray classes or cases involving multiple diseases, such as pneumonia and tuberculosis, further limiting its real-world applicability.

The study by Sharma and Guleria [21], which uses the Kermany dataset [29], and Patel P [38] employing VGG-16 with neural networks for pneumonia detection, presents several notable limitations. One key issue is the overlap in the datasets, as Patel P’s work already includes the Kermany dataset along with 576 additional COVID-19 images, leading to significant redundancy in pneumonia and normal lung images. This overlap undermines the validity of testing VGG-16’s efficiency on different datasets, as the model’s performance is likely influenced by the repetitive data. Furthermore, the computational demands of the VGG-16 architecture, which has approximately 138 million parameters, make it impractical for use in resource-constrained environments, hindering its real-world application. The study also suffers from a lack of class-based precision, recall, and F1 scores, which are critical for assessing model performance in medical diagnostics. These factors raise concerns about the model’s ability to deliver reliable, scalable results in clinical settings. To address these issues, future work would need to incorporate larger, more diverse datasets and optimize model architectures to reduce computational costs while improving classification accuracy and robustness.

Singh et al. [9] employed a Vision Transformer (ViT) model for pneumonia detection using the Kermany dataset [29]. Although the ViT model achieved high accuracy (97.61%), it suffers from limitations due to the relatively small Kermany dataset. While ViT excels in capturing global context and spatial relationships, its large computational cost and extended training time may not justify the performance gains over traditional CNNs, especially for smaller datasets. This highlights a critical limitation of the state-of-the-art approaches: for smaller datasets, large models like ViT may not provide substantial improvements, and simpler, more computationally efficient models may achieve similar results. This calls for careful consideration of dataset size, model complexity, and training cost in medical applications.

The ensemble method proposed by Kundu et al. [22] faces key limitations, including high computational costs due to combining three CNN models (GoogLeNet, ResNet-18, and DenseNet-121), making it unsuitable for low-resource settings. Its lower performance on the RSNA dataset [30] raises concerns about generalizability, while the handcrafted weighting mechanism risks overfitting to specific datasets, limiting adaptability to diverse imaging sources. Additionally, the lack of handling unknown or non-X-ray images reduces robustness, hindering the model’s performance on unseen or anomalous cases. These issues highlight the need for optimization to improve efficiency, handle non-X-ray classes, and enhance generalization across datasets and imaging devices.

The quaternion-based approach proposed by Singh and Tripathi [23] faces significant limitations, primarily due to its reliance on the relatively small and imbalanced Kermany dataset [29], which undermines its robustness and generalizability. Furthermore, the model’s accuracy and other key performance metrics lag state-of-the-art methods, limiting its applicability in real-world clinical settings. Addressing these challenges requires the use of larger, more diverse datasets and further optimization of the model to enhance its reliability and effectiveness.

## Section 3: Proposed Approach & Dataset

### 3.1) Explanation of This Approach

This study proposes a robust multi-class classification framework for chest X-ray (CXR) image analysis, categorizing images into four classes: normal, pneumonia, tuberculosis (TB), and non-X-ray. The inclusion of an “non-X-ray” class addresses a critical gap in previous studies by enabling the model to flag outlier or novel cases, improving robustness for real-world clinical applications where unexpected conditions may arise.

#### A. Dataset Compilation

To ensure diversity and broad representation of chest X-ray conditions, a comprehensive dataset was curated from multiple publicly available sources:

⍰ **Kermany et al. (2018)**: 38.23% (5856 images) [29].
⍰ **RSNA Dataset**: 22.85% (3500 images) [30].
⍰ **NIAID:** 22.84% (3499 images) [34].
⍰ **Shenzhen (4.32%, 662 images) & Montgomery (0.9%, 138 images)**: National Library of Medicine (NLM) datasets [35].
⍰ **Belarus:** 1.98% (304 images) [36].
⍰ **Non-X-ray Class Dataset (8.86%, 1357 images)**: Provided by Pavan Sanagapati, essential for identifying outlier cases [39].

This carefully curated dataset addresses limitations in prior studies that relied on smaller, less diverse datasets, ensuring better generalization across various imaging conditions and populations.

#### B. Model Architecture and Training

The framework employs fine-tuned **ResNet-18** and **DenseNet-121** architectures, both known for their ability to efficiently extract features from complex medical images. ResNet-18 leverages deep residual connections to mitigate vanishing gradient issues, while DenseNet-121 emphasizes feature reuse through densely connected layers. These architectures were selected due to their strong performance in medical imaging tasks.

Two distinct experimental setups were conducted:

⍰ **Without Class Weights**: Models were trained without applying a weighted loss function to address class imbalance.
⍰ **With Class Weights**: A weighted loss function was employed to ensure equitable learning across all categories, addressing the disparity in image availability among classes. The class weights applied were: Normal (0.7), Pneumonia (0.9), Non-X-ray (1.0), Tuberculosis (1.0).

#### C. Performance Evaluation

At the 24th epoch, the ResNet-18 model trained without class weights demonstrated exceptional class-wise metrics:

- **Normal**: Precision = 0.99, Recall = 0.98, F1-score = 0.98.
- **Pneumonia**: Precision = 0.98, Recall = 0.98, F1-score = 0.98.
- **Non-X-ray**: Precision = 1.000, Recall = 1.000, F1-score = 1.000.
- **Tuberculosis**: Precision = 1.0, Recall = 0.99, F1-score = 0.99.

The overall accuracy achieved was **98.77%**, with macro and weighted averages for precision, recall, and F1-scores reaching **0.99**. This highlights the robust performance of the ResNet-18 architecture even without the application of class weights.

### 3.2) Validity of This Approach

This approach exhibits outstanding performance while utilizing minimal computational resources. By incorporating diverse datasets, introducing the “non-X-ray” class, and employing weighted loss functions, it significantly enhances model robustness. These features make it highly suitable for real-world clinical deployment, particularly in resource-limited environments, delivering results that surpass expected benchmarks and even human-level accuracy.

### 3.3) Dataset description

This study utilizes multiple datasets comprising chest X-ray (CXR) images and non-X-ray images to develop and evaluate classification and detection models for pneumonia and tuberculosis (TB). The datasets include both publicly available collections and proprietary resources, summarized as follows:

#### OCT Dataset

The OCT dataset, derived from the Kermany dataset [29], contains 4,273 pneumonia-infected lung X-ray images and 1,583 normal lung X-ray images. These images are provided in high resolution, spanning a wide range of dimensions. The chest X-ray images (anterior-posterior views) were obtained from pediatric patients aged one to five years at the Guangzhou Women and Children’s Medical Center, Guangzhou. All imaging was conducted as part of the patients’ routine clinical care.

#### RSNA CXR Dataset

The RSNA pneumonia detection challenge dataset [30] includes approximately 30,000 chest X-ray images, of which 10,000 are normal, and the rest are abnormal with lung opacities. For this study, 3,500 TB-infected X-ray images were used, converted to Portable Network Graphics (PNG) format with a resolution of 512×512 pixels.

#### NIAID TB Dataset

The NIAID TB portal program dataset [34] contains 3,500 TB-positive CXR images (used 3,499) derived from seven different countries. All images are provided in Portable Network Graphics (PNG) format.

#### NLM Dataset

The National Library of Medicine (NLM) in the United States [35] has made two lung X-ray datasets publicly available:

- Montgomery County (MC) Database: Contains 138 posterior-anterior (PA) chest X-ray images, with resolutions of either 4,020×4,892 or 4,892×4,020 pixels. Among these, 57 images are from TB patients, while 80 images are from normal subjects.
- Shenzhen (CHN) Database: Comprises 662 PA chest X-ray images with a resolution of approximately 3,000×3,000 pixels. Of these, 336 images are from TB patients, and 326 images are from normal individuals.

Overall, the NLM database includes 406 normal and 393 TB-infected X-ray images.

#### Belarus Dataset

The Belarus dataset [36] was collected as part of a drug resistance study conducted by the National Institute of Allergy and Infectious Diseases (NIAID) in collaboration with the Ministry of Health, Republic of Belarus. It consists of 304 TB-infected CXR images from 169 patients. The chest radiographs were captured using the Kodak Point-of-Care 260 system, with an image resolution of 512×512 pixels.

#### Non-X-ray or Unknown Dataset

This dataset contains images of non-X-ray objects, such as bikes, cars, cats, dogs, flowers, horses, and humans. It is intended for tasks involving identification of *non-X-ray* labels, making it valuable for multi-class classification and open-set recognition challenges. Containing 1357images. This dataset provides sufficient diversity to act as an outlier group, distinguishing X-ray images from non-X-ray images effectively [39].

### 3.4) Custom Dataset and Splits

To evaluate the performance of the models, a custom dataset was created by organizing images into three distinct folders: train, validation, and test. Simultaneously, a CSV mapping file was generated to ensure data transparency by recording the image name, label, source, and split for every sample. This structured approach facilitates proper evaluation, maintains transparency, and reduces the risk of overfitting during model training.

The dataset distribution is as follows:

- **Training Set:** 4667 Normal, 3633 Pneumonia, 3573 Tuberculosis, 1155 Unknown images.
- **Validation Set:** 274 Normal, 213 Pneumonia, 207 Tuberculosis, 67 Non-X-ray images.
- **Testing Set:** 584 Normal, 427 Pneumonia, 417 Tuberculosis, 135 Non-X-ray images. The split follows these proportions:
- **Training Set:** 85% from each specific source.
- **Validation Set:** 5% of each specific source.
- **Testing Set:** 10% of each specific source.

#### Preserving Proportional Representation

This proportional splitting method is particularly important for datasets with fewer samples, such as the NLM (Montgomery and Shenzhen) and Belarus datasets. Without this careful allocation, smaller datasets could have been excluded entirely from certain splits due to random selection during mixing. By enforcing proportional splits, the method guarantees:

1. **Balanced Contribution:** Each dataset, regardless of size, contributes meaningfully to the training, validation, and testing processes.
2. **Fair Evaluation:** The testing set reflects the performance of the model across all datasets, ensuring that results are not skewed toward larger datasets.
3. **Generalization:** By exposing the model to data from all sources in both training and evaluation, it can generalize better across diverse datasets without favoring specific ones.

## Section 4: Methodology and Model Configuration

### 4.1) Preprocessing

To prepare the chest X-ray images for both training and evaluation, a series of preprocessing steps were applied to ensure compatibility with the ResNet-18 and DenseNet-121 architectures. The preprocessing pipeline was implemented using the **torchvision** library, with specific transformations for the training and validation datasets:

#### A. Training Data Preprocessing

The training dataset underwent various augmentations to increase the diversity of input samples and improve the model’s robustness. The following transformations were applied:

1. **Resizing:** All images were resized to a uniform size of 224×224 pixels to meet the input size requirements for both ResNet-18 and DenseNet-121 architectures.
2. **Grayscale Conversion to RGB:** Since the chest X-ray images were initially in grayscale, they were converted to RGB format by duplicating the single grayscale channel across the three-color channels. This ensures compatibility with the pre-trained ResNet-18 and DenseNet-121 models, which expect RGB inputs.
3. **Random Horizontal Flip:** To simulate natural variations in X-ray orientations, a random horizontal flip was applied to the images during training. This augmentation helped the model generalize better by exposing it to more variations of the X-ray images.
4. **Random Rotation:** Small random rotations (between −5 and +5 degrees) were applied to account for minor positional variations in X-ray scans, further enhancing the model’s ability to handle different orientations and perspectives.
5. **Conversion to Tensor:** The images were converted into PyTorch tensors, which is the required format for input into the deep learning models. This transformation is essential for the models to perform matrix operations during training.
6. **Normalization:** To align the pixel intensity distribution with the ImageNet Dataset, the images were normalized using specific mean and standard deviation values:
7. Mean: [0.485, 0.456, 0.406]
8. Standard deviation: [0.229, 0.224, 0.225]

#### B. Validation and Testing Data Preprocessing

To evaluate the model’s performance without introducing data variability, a minimal preprocessing approach was applied to the validation and testing datasets. The transformations applied were:

1. **Resizing:** Images were resized to 224×224 pixels to match the input size of the models.
2. **Grayscale Conversion to RGB:** Similar to the training data, grayscale images were converted to RGB format to ensure consistency in input across all stages of model training and evaluation.
3. **Conversion to Tensor:** Like the training images, the validation and test images were converted into PyTorch tensors.
4. **Normalization:** Validation and test images were normalized using the same mean and standard deviation values as the training data, ensuring that the evaluation process reflects the preprocessing done during training.

#### C. Custom Dataset Class

To handle the image dataset efficiently, a custom Dataset class was created. This class allows for seamless loading and transformation of the image data, ensuring that the appropriate preprocessing steps are applied to each image during training and evaluation.

The dataset class, which accepts a dataframe object containing the file paths and corresponding labels for each image, applies the required transformations as specified in the code. The images are loaded and transformed before being passed into the model for training or evaluation.

#### D. Cross-validation for Redundancy Detection

To ensure data integrity and eliminate redundancies, a duplicate detection algorithm was implemented using image hashing techniques.

1. **Normalization**: Images were resized to 100×100 pixels and converted to grayscale for uniformity.
2. **Hashing**: A unique hash was generated for each image using the MD5 algorithm.
3. **Comparison**: Duplicates were identified by comparing hash values, ensuring robustness against format (JPEG, PNG, BMP) and resolution variations.

### 4.2) System Configuration

#### Hardware

- **Laptop:** NVIDIA RTX 3060, AMD Ryzen 7 5800H
- **RAM:** 24 GB DDR4 3200 MHz
- **Storage:** 100 GB disk drive (SSD)
- **Operating System:** Windows 11

#### Software

- **IDE:** Visual Studio Code with Jupyter Notebook extension
- **Environment:** Conda, Python 3.11
- **Key Libraries:** PyTorch, NumPy, Pandas, Matplotlib, TorchVision, Scikit-learn, Pillow

This setup provided the necessary computational resources and software tools as well as freedom for model training and evaluation.

### 4.3) Model Architecture and Configuration

This section elaborates on the selection and customization of the ResNet-18 and DenseNet-121 architectures, detailing their features, specific modifications, and rationale for use in this study.

#### A. Model Selection ResNet-18

ResNet-18, introduced by He et al. [40], is a highly efficient deep learning model built on the residual learning framework. The use of residual blocks in ResNet overcomes the vanishing gradient problem, which is a common challenge in training deep neural networks. Unlike traditional convolutional networks with monotonically progressive mappings, ResNet-18 employs **skip connections** that perform identity mapping, allowing gradients to flow directly across layers. This mechanism significantly improves network optimization and model accuracy without adding extra parameters or computational overhead.

ResNet-18 has **11.4 million trainable parameters**, making it computationally lightweight and suitable for environments with limited resources. Initially trained on the ImageNet dataset, ResNet-18 has demonstrated strong generalization capabilities and is particularly effective for tasks requiring high accuracy in resource-constrained settings.

#### DenseNet-121

DenseNet-121, proposed by Huang et al. [41], offers a unique approach to feature extraction by leveraging dense connectivity. In DenseNet, each layer receives input not only from the previous layer but also from **all preceding layers**, creating a direct connection between layers. This results in efficient feature reuse and significantly enhances feature representation. Unlike ResNet, which adds the output of residual layers, DenseNet concatenates feature maps from previous layers with those of the current layer.

This architecture reduces the number of redundant computations by limiting the number of channels in convolutional layers. Despite having **only 7.4 million parameters**, DenseNet-121 maintains a rich feature extraction capability while being memory-efficient. Its concatenation-based design diminishes redundancy, making it a practical choice for real-world applications. Although DenseNet-121 has fewer parameters than ResNet-18, its dense connections can introduce computational overhead, leading to longer training times. However, the model compensates for this with enhanced accuracy and efficiency in feature learning, especially for complex datasets.

#### B. Model Configuration

Both ResNet-18 and DenseNet-121 were customized and configured for the specific requirements of this study, with attention to compatibility with the dataset and the research objectives:

1. Input Layer: Both architectures were adapted to accept input images resized to dimensions of **224×224×3**. This matches the preprocessed chest X-ray images, which were converted to RGB to meet model requirements.
2. Output Layer: The original classification heads were replaced to accommodate the **four target classes**: **Normal, Pneumonia, Tuberculosis, Unknown/Non-X-ray**.

- **ResNet-18**: The original fully connected (FC) layer was replaced with a custom FC layer with 4 output nodes for multi-class classification.
- **DenseNet-121**: The final classifier block was modified to include a fully connected layer with 4 output nodes.

### 4.4) Model Training and validation

The training process for both ResNet-18 and DenseNet-121 involved careful configuration and systematic monitoring to optimize their performance in classifying chest X-ray images across four categories: Normal, Pneumonia, Tuberculosis, and Unknown/Non-X-ray. The models were trained with class-specific adjustments to handle the dataset’s inherent imbalance, and TensorBoard was employed to monitor key performance metrics throughout the training process.

#### A. Training Setup and Procedure

- **Epochs**: ResNet-18 was trained for 60 epochs, while DenseNet-121 was trained for 120 epochs due to its longer convergence time. These settings were selected based on the computational constraints and the models’ individual requirements for sufficient training.
- **Learning Rate**: A starting learning rate of 0.0001 was employed for both models, with a decay schedule to gradually reduce the learning rate over time, helping to achieve better convergence and avoid overfitting.
- Loss Function:

- **Weighted Cross-Entropy Loss**: To address the class imbalance inherent in the dataset, class-specific weights were computed and applied. The weights were determined based on the inverse frequency of each class, as discussed in Section 5.1 (Handling Class Imbalance).
- **Non-Weighted Cross-Entropy Loss**: For comparative analysis, models were also trained without class weighting to evaluate the impact of handling class imbalance on performance metrics.
- **Optimizer**: The Adam optimizer was selected for its effectiveness in gradient-based optimization, which was essential for training deep convolutional neural networks efficiently. Its adaptive learning rate capabilities ensured that both models could learn effectively without extensive tuning.

#### B. Performance Metrics and Logging

To comprehensively monitor training progress and model performance, the following metrics were logged at the end of each epoch:

1. **Training Loss**: This metric indicated how well the model was fitting to the training data.
2. **Training Accuracy**: This reflected the model’s ability to make correct predictions on the training dataset, providing a measure of generalization during training.
3. **Validation Loss**: This indicated how well the model was generalizing to unseen data, calculated using the validation dataset.
4. **Validation Accuracy**: This measured the model’s performance on the validation dataset, providing an additional gauge of its generalization capability.
5. **Epoch Time**: The total time taken to complete one full training cycle.
6. **Validation Time**: The time spent evaluating the model on the validation dataset after each epoch.

To track training and evaluation progress more effectively, TensorBoard was used to visualize the training and validation losses, accuracies, and time spent per epoch. This not only provided insights into model performance but also helped in adjusting training parameters and identifying potential issues such as overfitting or underfitting.

#### C. Model Evaluation and Saving

At the end of each epoch, the model weights were saved to ensure that the best-performing model could be easily identified. This also allowed for further analysis and comparison of model performance over time. The training loop involved performing the forward pass, calculating the loss, backpropagating the error, and updating the model weights. In parallel, the models were evaluated on the validation set after each epoch to monitor how well they generalized to unseen data.

#### D. Impact of Class Weights

Class imbalance is a common challenge in machine learning, particularly in medical image classification, where models often perform well on majority classes but poorly on minority ones. To address this, various techniques such as class-specific data augmentation or synthetic data generation can be used. In this study, however, the best results were achieved by adjusting the class weights during training.

Class weights were computed based on the inverse frequency of each class, helping the model focus more on the underrepresented classes. The weight for each class iii was calculated using the formula:

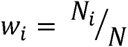

Where:

- N is the total number of samples.
- N_l_ is the number of samples in each class

This formula assigns higher weights to underrepresented classes. The class distributions in this study from section-3.2, the calculated class weights were as follows: Normal class (0.7), Pneumonia class (0.9), Tuberculosis class (0.9), and Unknown/non-X-ray class (2.8).

Based on these values and their clinical relevance, the weights were adjusted as follows:

- **Normal class (weight 0.7)**: Due to the largest number of samples (4,667), a lower weight was applied to prevent overemphasizing this class.
- **Pneumonia class (weight 0.9)**: With 3,633 images, a higher weight was assigned to ensure adequate attention to this clinically significant class.
- **Tuberculosis class (weight 1.0)**: Despite a similar number of images as pneumonia (3,573), tuberculosis was assigned a weight of 1.0 to maintain balance across the primary lung conditions.
- **Unknown and non-X-ray class (weight 1.0)**: The 1,155 samples of diverse images (e.g., cats, dogs, flowers, humans) were assigned a weight of 1.0 to help the model distinguish these outliers effectively.

Both models were trained with and without class weights. By training both models with these weighted settings, the study ensured balanced performance across all classes, which was crucial for handling the class imbalance problem. The results from training with and without class weights were compared to assess the impact of class weighting on model accuracy, robustness, and generalization.

This approach, combining optimized model configurations, monitoring tools, and class-weight adjustments, ensured that both ResNet-18 and DenseNet-121 models were effectively trained and evaluated for their performance on chest X-ray image classification tasks.

### 4.5 Model Testing

The evaluation of the trained models (ResNet-18 and DenseNet-121) was performed using a test dataset consisting of four distinct classes: Normal, Pneumonia, Tuberculosis, and Unknown/Non-X-ray. The testing phase aimed to assess the model’s ability to generalize to unseen data and provide robust classification performance.

#### A. Testing Setup

The testing pipeline began with the loading of pre-trained model weights, where the models were set to evaluation mode to ensure no gradient calculations occurred during inference. The models were then placed on the available computing device (CPU or GPU), allowing for efficient computation.

For both models, the following preprocessing steps were applied to the test dataset to ensure consistency with the training data:

- **Resizing**: Images were resized to 224×224 pixels to meet the input size requirements of the models.
- **Grayscale to RGB Conversion**: The grayscale chest X-ray images were converted into RGB format to align with the input expectations of the ResNet-18 model.
- **Normalization**: Images were normalized using the same mean and standard deviation values as during training, ensuring the distribution of pixel intensities aligned with the model’s expectations.

#### B. Testing Procedure

For each of the four classes (Normal, Pneumonia, Tuberculosis, and Unknown/Non-X-ray), a loop iterated over all test images. Each image was passed through the model to generate predictions. The predicted labels were compared against the ground truth labels, allowing for the calculation of several evaluation metrics.

The following key performance indicators were logged and computed during the testing phase:

1. **Accuracy**: This was the percentage of correctly predicted images across all test samples. The accuracy is a fundamental metric for assessing the overall performance of the model in classifying chest X-ray images.
2. **Classification Report**: This detailed report provided metrics such as Precision, Recall, F1-score, and Support for each class. These metrics provided a deeper insight into the model’s performance, particularly with respect to how well it identified each class in the imbalanced dataset.
3. **Confusion Matrix**: A confusion matrix was generated to visualize the distribution of true positive, true negative, false positive, and false negative predictions for each class. This matrix enabled a detailed evaluation of misclassifications and helped identify any patterns or areas of weakness in the model’s performance.
4. **Per-Class Performance**: Each class (Normal, Pneumonia, Tuberculosis, Non-X-ray) was evaluated separately, allowing for a comparison of model performance across different categories. This was particularly important for addressing the class imbalance in the dataset.
5. **Grad**-**CAM**: To visualize efficiency and accuracy of the model we take last layer and visualize the region which contributes most in models’ detection.

## Section 5: Results and Evaluation

### 5.1) Metrics overview

#### A. Definitions of Metrics

1. **True Positives (TP)**: Cases where the model correctly predicts the actual class.

1. Example: If an X-ray labeled as “Pneumonia” is predicted as “Pneumonia,” it counts as a TP.
2. **False Positives (FP)**: Cases where the model incorrectly predicts a given class.

1. Example: An X-ray labeled as “Normal” but predicted as “Pneumonia” is a FP.
3. **True Negatives (TN)**: Cases where the model correctly predicts the absence of a given class.

1. Example: If the model predicts “Normal” and the actual label is also “Normal,” it counts as a TN.
4. **False Negatives (FN)**: Cases where the model fails to predict the actual class.

1. Example: An X-ray labeled as “Pneumonia” but predicted as “Normal” is a FN.

#### B. Metrics Formulas

- **Accuracy**: Measures the overall correctness of the model’s predictions.

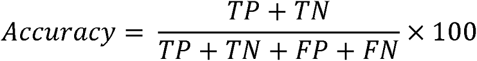
- **Precision**: Proportion of positive predictions that are correct, minimizing False Positives (FP).

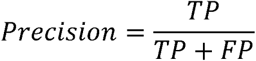
- **Recall (Sensitivity)**: Measures the ability to correctly identify all instances of a given class, minimizing False Negatives (FN).

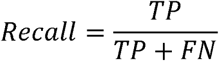

**F1 Score**: Balances precision and recall, especially useful for imbalanced datasets.

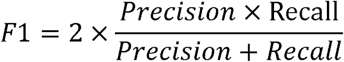

### 5.2) Training results

The training results for ResNet-18 and DenseNet-121 were carefully analyzed to evaluate the impact of class weights and identify optimal configurations for effective classification. Metrics such as accuracy, loss, and the epoch of peak performance were tracked and compared under two scenarios: training with and without class weights. TensorBoard was employed to log the progress and visualize the performance trends throughout the training process.

Below in Table 1 we present a summary of (Figure 18 – 25) the training outcomes. The table shows the best-performing models in terms of training results. Specifically, two models were selected for their superior performance on the test set, while two others excelled in training performance compiling 8 models. Both scenarios—training with and without class weights—are included for comparison.

**Table 1.** Best Training Summery.

| Model Name | Weighted Loss | Train Accuracy (%) | Train Loss | Epoch | Best on | Time (hr) | Validation Accuracy (%) | Validation Loss |
| --- | --- | --- | --- | --- | --- | --- | --- | --- |
| ResNet18 | No | 99.34 | 0.0193 | 25 | Test | 1.66 | 98.69 | 0.0896 |
|  | Yes | 99.65 | 0.0090 | 37 | Test | 2.62 | 98.42 | 0.0620 |
|  | No | 99.84 | 0.0070 | 39 | Train | 2.55 | 98.69 | 0.0798 |
|  | Yes | 99.69 | 0.0076 | 52 | Train | 3.67 | 98.42 | 0.0714 |
| DenseNet-121 | No | 99.76 | 0.0096 | 120 | Test, Train | 9.88 | 97.90 | 0.0741 |
|  | Yes | 99.59 | 0.0105 | 108 | Test, Train | 10.63 | 97.63 | 0.0105 |

#### A. ResNet-18 Performance Analysis

- **Without Class Weights:**

- At **epoch 39**, the model achieved its highest training accuracy of **99.84%**, with a corresponding loss of **0.0070**. However from Table 2, this configuration showed slight overfitting, as its test accuracy dropped to **97.71%**, highlighting a lack of generalization compared to earlier epochs.
- At **epoch 25**, the model delivered the best overall performance, achieving a test accuracy of **98.75%**, with robust generalization to unseen data.
- With Class Weights:

- At **epoch 52**, the model attained a training accuracy of **99.69%**, but test accuracy decreased slightly to **98.03%**, indicating that the longer training process may have contributed to marginal overfitting.
- At **epoch 37**, the model achieved a test accuracy of **98.49%**, maintaining strong performance across all classes, though it underperformed slightly compared to the non-weighted configuration at epoch 25.

#### B. DenseNet-121 Performance Analysis

- **Without Class Weights:**

- At **epoch 120**, the model achieved a training accuracy of **99.76%** and a test accuracy of **97.97%**, demonstrating stable performance across both training and testing phases.
- With Class Weights:
- At **epoch 111**, the model achieved a slightly lower test accuracy of **97.71%**, comparable to its performance without class weights. While training accuracy was consistent, the results did not significantly improve the generalization capacity of the model.

#### C. Overview

The addition of class weights increased training time slightly but did not provide significant advantages in terms of accuracy or loss. Models trained without class weights exhibited better generalization and slightly higher test accuracy, suggesting that the dataset did not benefit substantially from reweighting underrepresented classes.

**Table 2.** Best Test Summery.

| Model Name | Optimizer Weight | Model | Acc % | Pre | F1 | N-Rec | N-Pre | P-Rec | P-Pre | T-Rec | T-Pre | U-Rec | U-Pre | Best On |
| --- | --- | --- | --- | --- | --- | --- | --- | --- | --- | --- | --- | --- | --- | --- |
| ResNet18 | Yes | 37 | 98.49 | 0.98 | 0.98 | 0.97 | 0.99 | 0.99 | 0.97 | 0.99 | 0.99 | 1.00 | 1.00 | Test |
|  | Yes | 52 | 98.03 | 0.98 | 0.98 | 0.97 | 0.97 | 0.98 | 0.98 | 0.98 | 0.99 | 1.00 | 1.00 | Train |
|  | No | 25 | 98.75 | 0.99 | 0.99 | 0.98 | 0.99 | 0.99 | 0.98 | 0.99 | 1.00 | 1.00 | 1.00 | Test |
|  | No | 39 | 97.71 | 0.98 | 0.98 | 0.96 | 0.97 | 0.97 | 0.99 | 0.99 | 0.97 | 1.00 | 1.00 | Train |
| DenseNet-121 | Yes | 111 | 97.71 | 0.98 | 0.98 | 0.96 | 0.98 | 0.98 | 0.98 | 0.99 | 0.96 | 1.00 | 1.00 | Train, Test |
|  | No | 120 | 97.97 | 0.98 | 0.98 | 0.97 | 0.97 | 0.98 | 0.98 | 0.98 | 0.99 | 1.00 | 0.99 | Train, Test |

### 5.3) Test Result

The final evaluation of ResNet-18 and DenseNet-121 models on the test set was conducted using key metrics: accuracy, precision, recall, and F1 score. These metrics were calculated across four classes: Normal, Pneumonia, Tuberculosis, and Non-X-ray. Metrics for each class (e.g., N-Recall for Normal Recall, P-Precision for Pneumonia Precision) were also included to ensure a thorough assessment of classification performance on Table 2, Figure 26,27.

#### A. ResNet-18 Performance Analysis

- **Without Class Weights:**

- **Epoch 25 (Best on Test):** The model achieved the highest test accuracy of 98.75%, indicating strong generalization to unseen data.
- **Epoch 39 (Best on Train):** While training accuracy reached a high of 99.84%, this model exhibited slight overfitting, as evidenced by lower test accuracy (97.71%) compared to the optimal epoch 25 performance.
- With Class Weights:

- **Epoch 37 (Best on Test):** Achieved a test accuracy of 98.49%, maintaining excellent performance across all classes, though slightly lower than the model without class weights at epoch 25.
- **Epoch 52 (Best on Train):** The model demonstrated a training accuracy of 99.69%, with a slightly lower test accuracy of 98.03%. Despite the reduced test accuracy, performance was comparable to the test-optimal epoch 37.

#### B. DenseNet-121 Performance Analysis

- **Without Class Weights:**

- **Epoch 120 (Best on Test and Train):** The model achieved a test accuracy of 97.97%, slightly outperforming its class-weighted counterpart.
- With Class Weights:
- **Epoch 111 (Best on Test and Train):** The model displayed consistent performance across both the training and testing phases, achieving an accuracy of 97.71%, which is comparable to the best performance seen without class weights at epoch 107.

#### C. Key Observations

- **Impact of Class Weights:** The addition of class weights had a minimal effect on the test set accuracy and performance metrics. Models trained without class weights generally achieved slightly higher test accuracy and demonstrated better generalization.
- Comparison: ResNet-18 vs. DenseNet-121:

- ResNet-18 consistently outperformed DenseNet-121 in test accuracy, particularly in models trained without class weights.
- DenseNet-121, however, showed robust performance across both training and testing phases, with less variation in performance metrics.
- Performance Across Classes:

- Both models excelled in classifying the “non-X-ray” category, achieving near-perfect recall and precision.
- Minor performance discrepancies were observed in the “Normal” and “Pneumonia” classes, where recall occasionally lagged behind precision.
- Generalization: Models trained without class weights consistently demonstrated superior generalization, suggesting that the application of class weights did not provide significant benefits for this dataset.

### 5.4) Test Visualization

To comprehensively evaluate the performance of ResNet-18 and DenseNet-121, confusion matrices were generated for the test set. These matrices offer a clear view of classification performance across the four classes: Normal, Pneumonia, Tuberculosis, and Non-X-ray. Diagonal entries indicate correct classifications, while off-diagonal values represent misclassifications. Additionally, Grad-CAM visualizations were employed to highlight the regions of input images that contributed most significantly to the models’ predictions, facilitating interpretability and validating their clinical relevance.

#### A. Confusion Matrix Analysis for ResNet-18 (Best Test Model)

The ResNet-18 model without optimizer weights (Epoch 25) demonstrated outstanding test performance, achieving high classification accuracy across all classes. Below at Figure 1 are the observations:

1. Normal Class

- Correctly classified **537 samples** as “Normal.”
- Misclassified **9 samples** as “Pneumonia” and **2 samples** as “Tuberculosis.”
2. Pneumonia Class

- Correctly classified **422 samples** as “Pneumonia.”
- Misclassified **5 samples** as “Normal.”
3. Non-X-ray Class

- Achieved perfect classification, with all **135 samples** correctly identified as “Non-X-ray.”
4. Tuberculosis Class

- Correctly classified **414 samples** as “Tuberculosis.”
- Misclassified **3 samples** as “Normal.”

**Figure 1.**
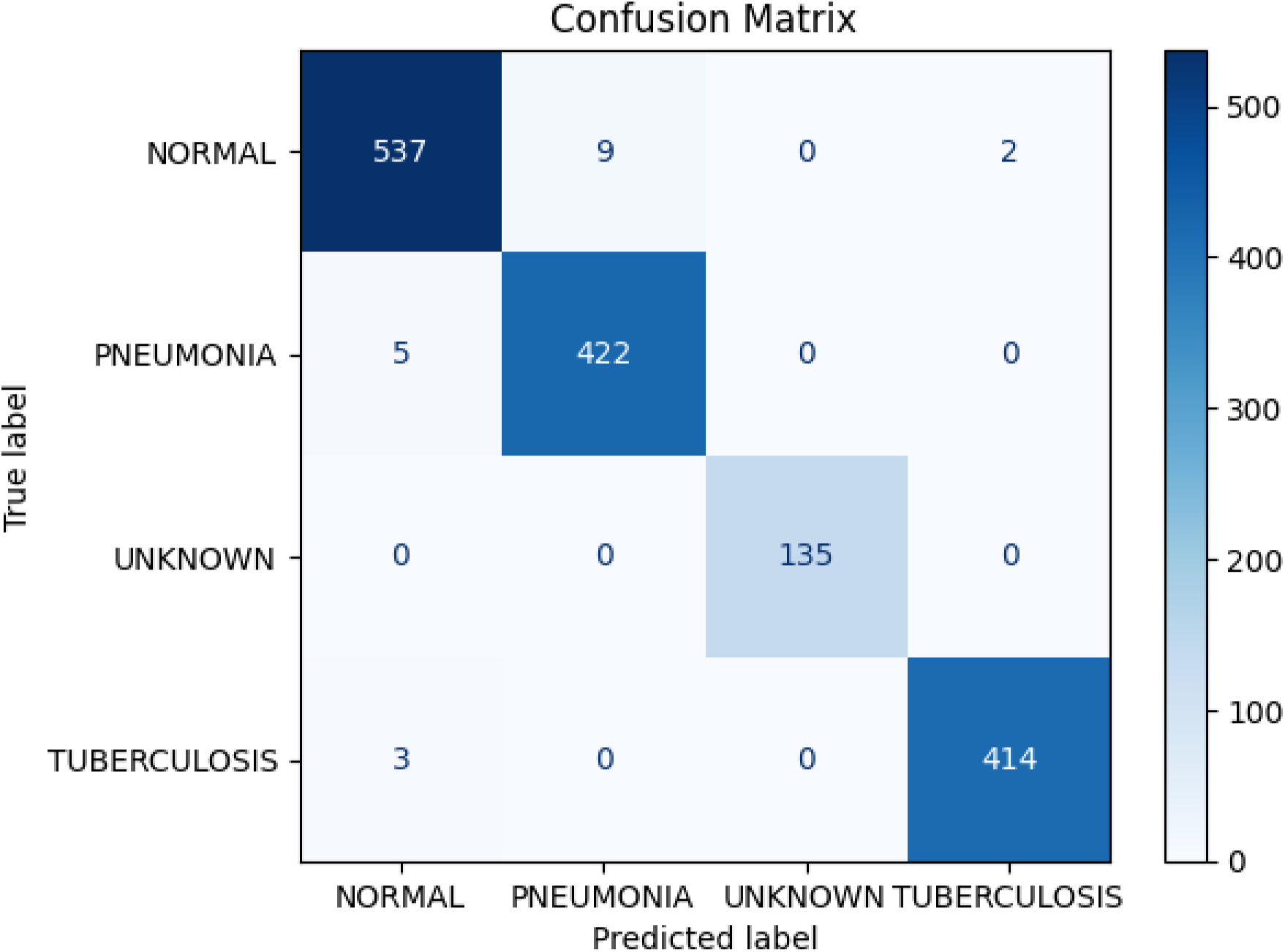
Confusion Metrics Resnet18

#### B. Confusion Matrix Analysis for DenseNet-121 (Best Test Model)

The DenseNet-121 model without weighted loss function (Epoch 120) also exhibited strong performance, with slightly more misclassifications in certain classes compared to ResNet-18. Observations include in Figure 2:

1. Normal Class

- Correctly classified **533 samples** as “Normal.”
- Misclassified **7 samples** as “Pneumonia” and **8 samples** as “Tuberculosis.”
2. Pneumonia Class

- Correctly classified **418 samples** as “Pneumonia.”
- Misclassified **9 samples** as “Normal.”
3. Non-X-ray Class

- Achieved perfect classification, with all **135 samples** correctly identified as “non-X-ray.”
4. Tuberculosis Class

- Correctly classified **410 samples** as “Tuberculosis.”
- Misclassified **6 samples** as “Normal” and **1 sample** as “Non-X-ray.”

**Figure 2.**
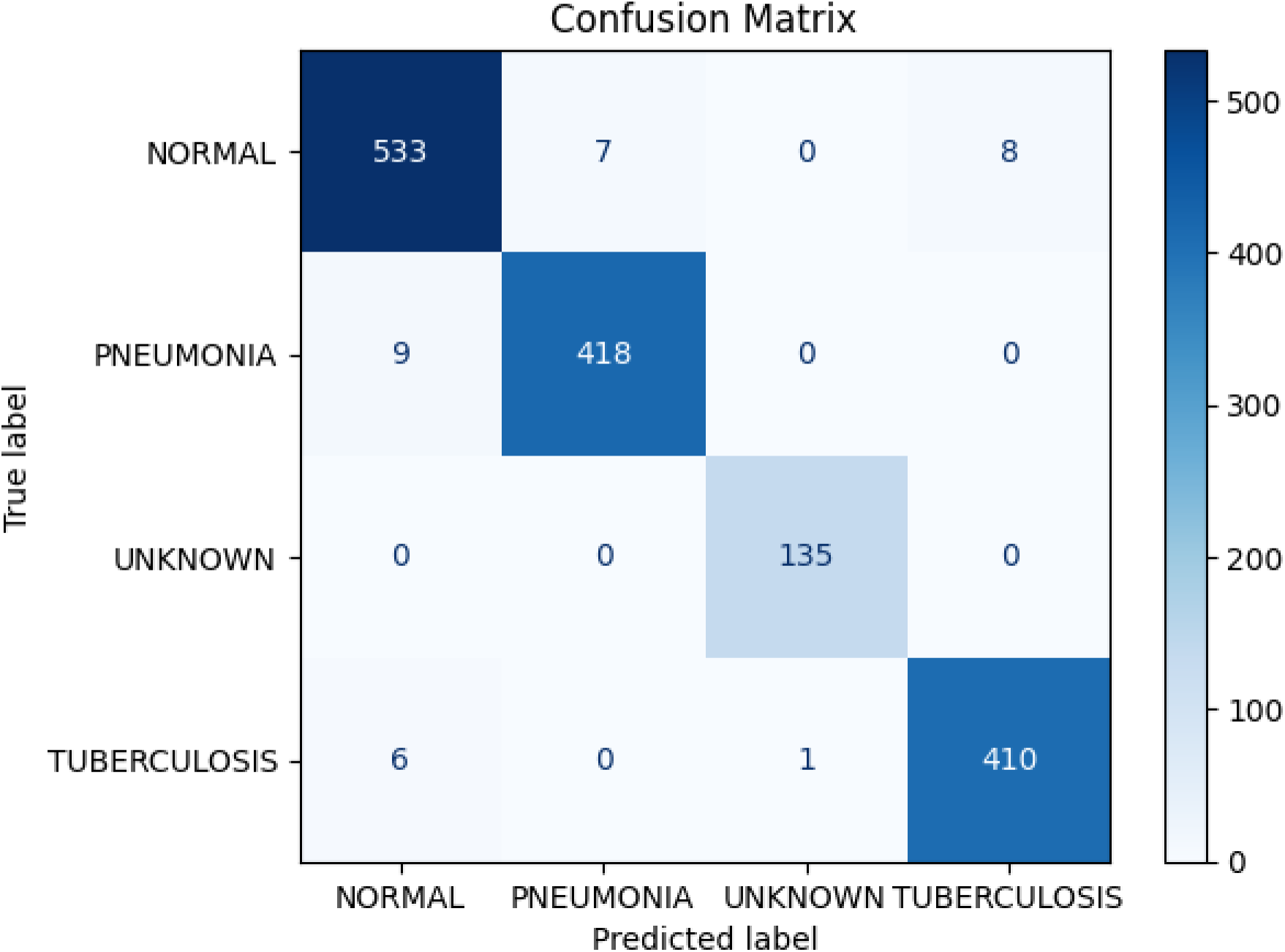
Confusion Metrics DenseNet-121

#### C. Grad-CAM Visualizations

Grad-CAM visualizations were employed to gain insights into the decision-making processes of the ResNet-18 and DenseNet-121 models. These heatmaps illustrate the areas of the input image that contributed most to the model’s predictions, enhancing interpretability and validating the models’ reliability for clinical applications.

#### ResNet-18 Grad-CAM Analysis

To analyze the model’s predictions, 7 visualizations were created:

- **4 Correct Predictions**: One from each class (Normal: Figure 3, Pneumonia: Figure 5, Tuberculosis: Figure 7, Non-X-ray: Figure 9).
- **3 Incorrect Predictions**: One misclassified example per class (Normal: Figure 4, Pneumonia: Figure 6, Tuberculosis: Figure 8). No misclassifications occurred in the Non-X-ray class.

**Figure 3.**
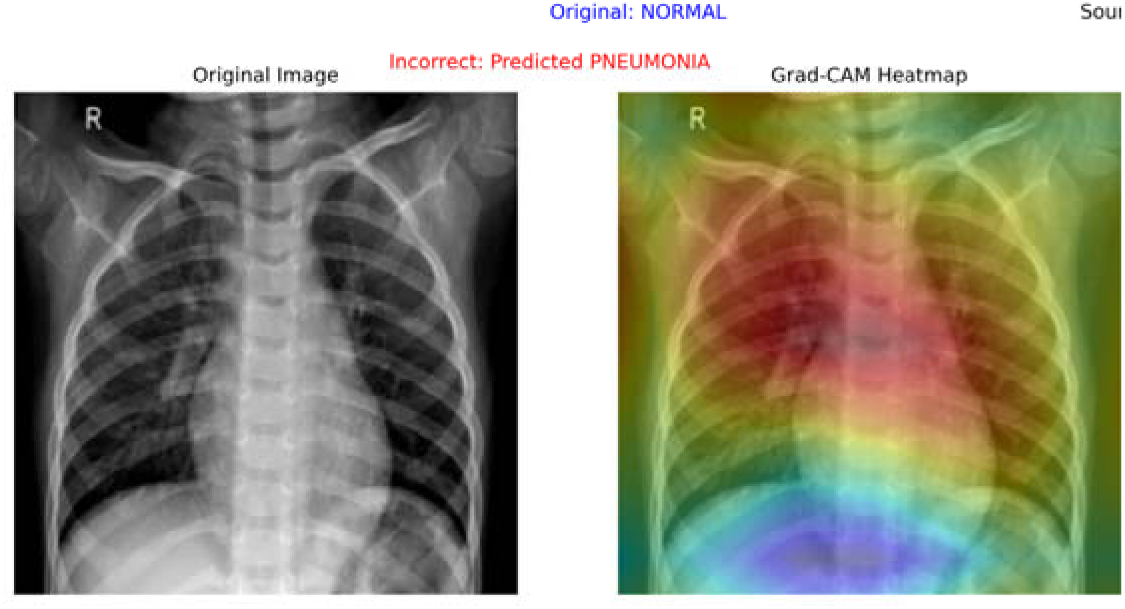
Incorrect Pneumonia Prediction of Normal

**Figure 4.**
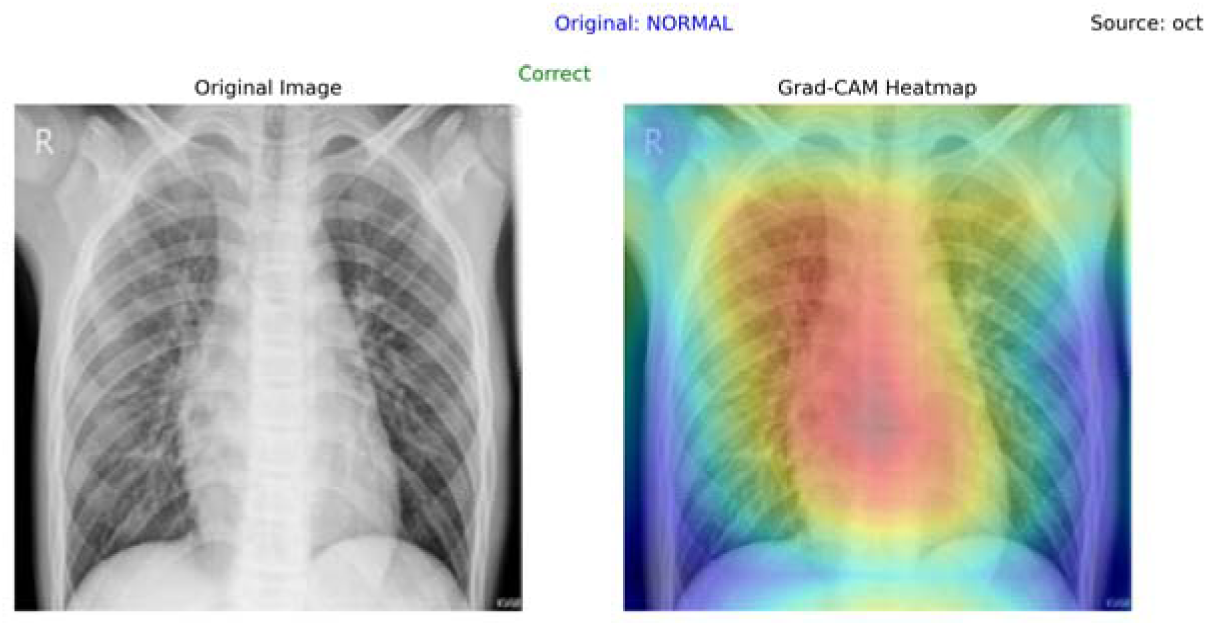
Correct Normal Prediction

**Figure 5.**
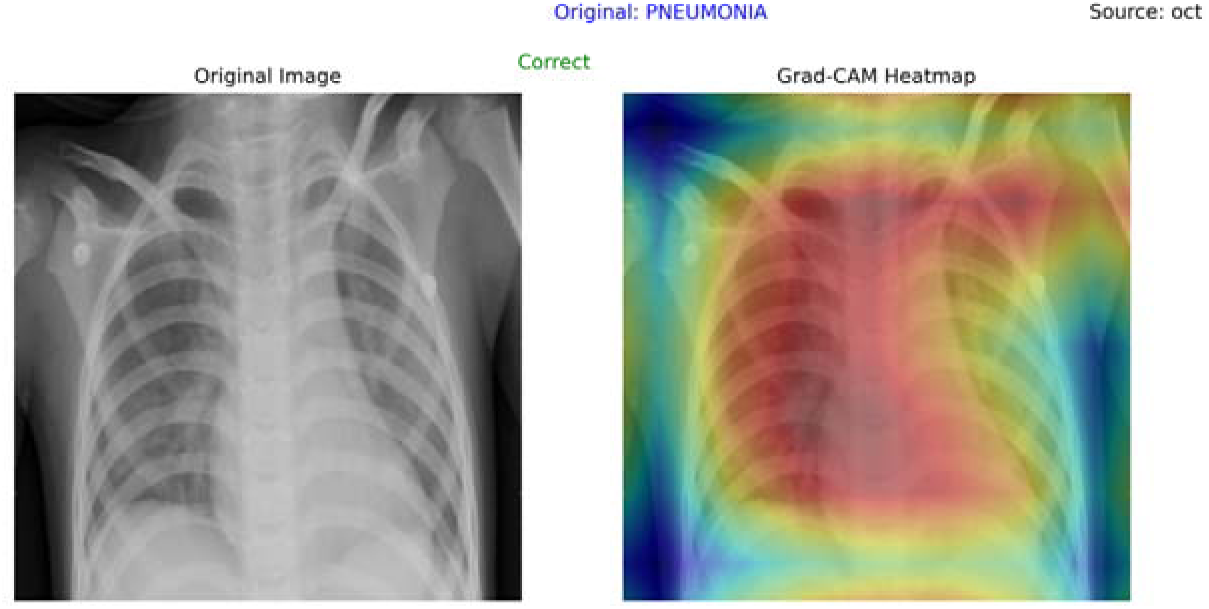
Correct Pneumonia Prediction

**Figure 6.**
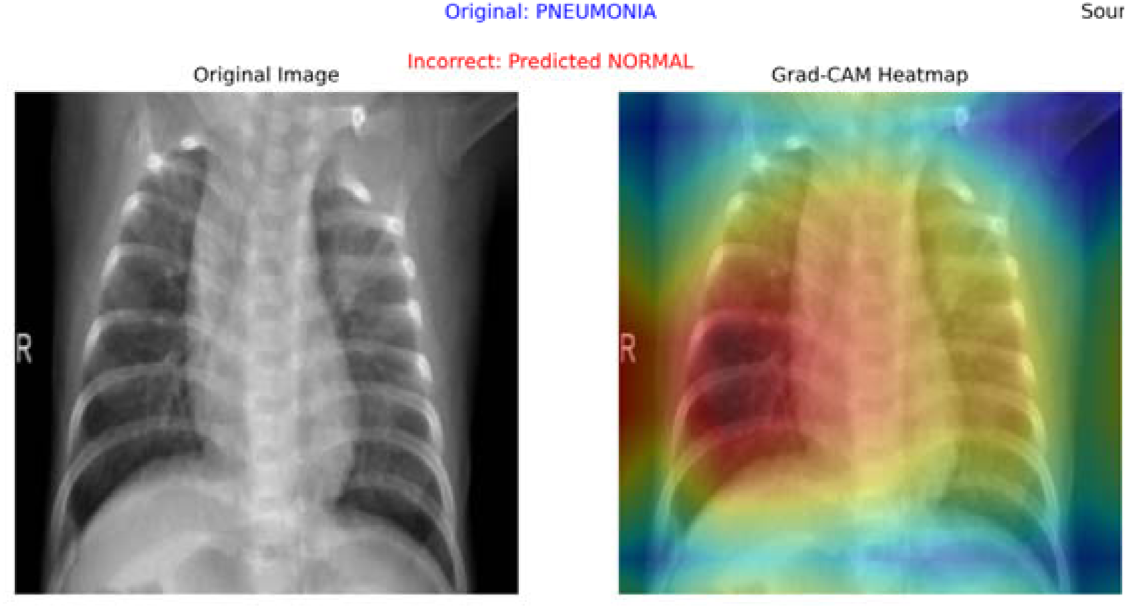
Incorrect Normal Prediction of Pneumonia

**Figure 7.**
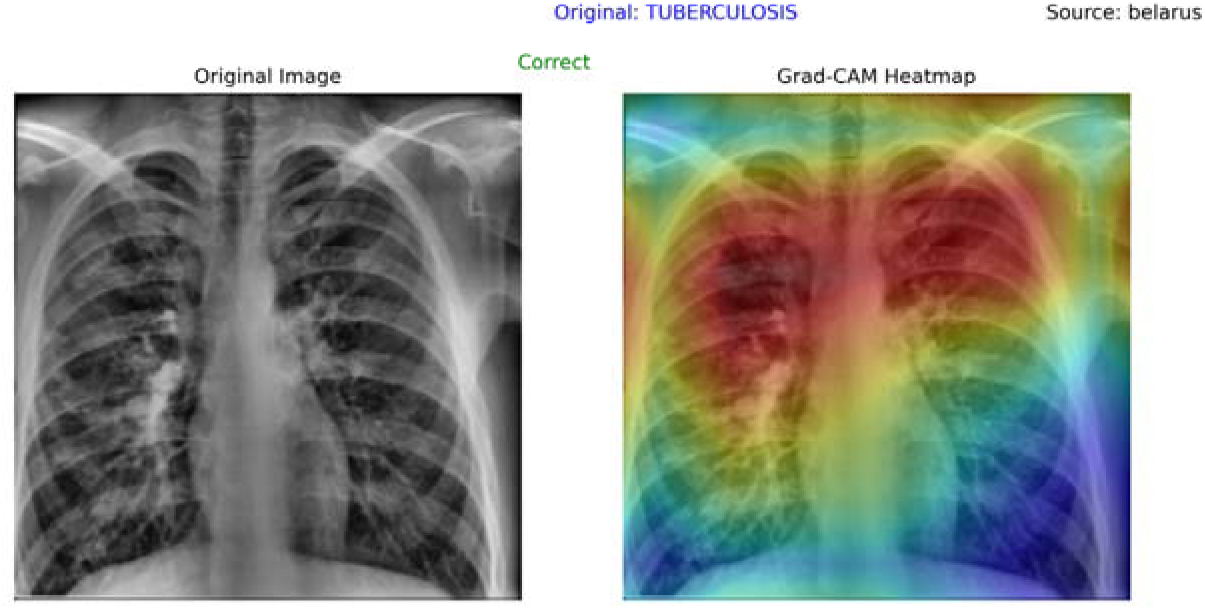
Correct Tuberculosis Prediction

**Figure 8.**
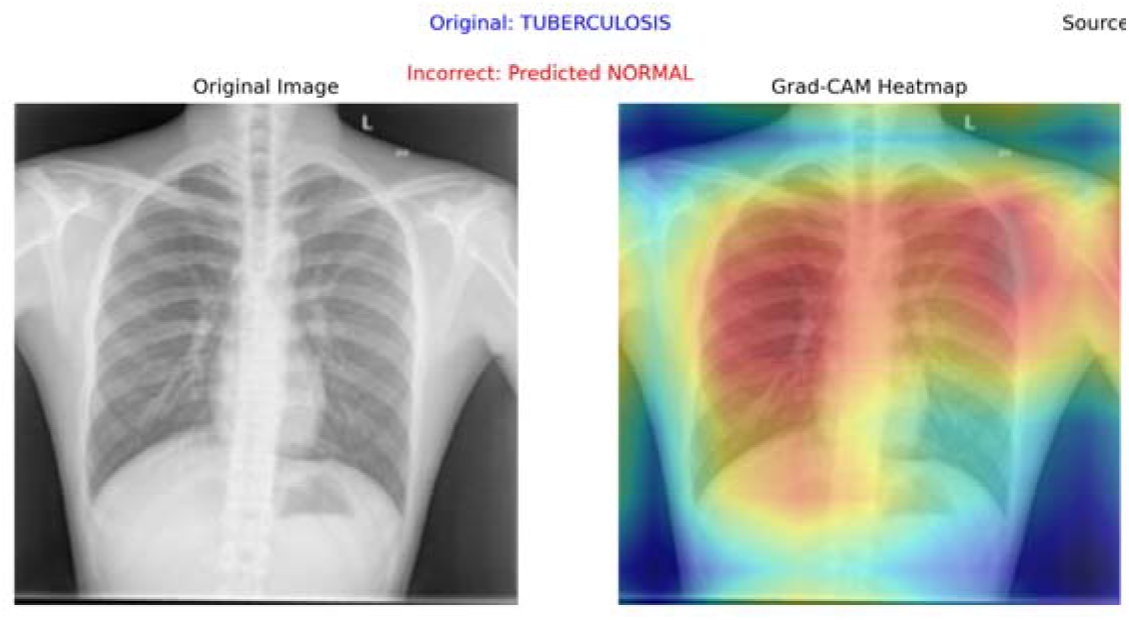
Incorrect Normal Prediction of Tuber ulosis

**Figure 9.**
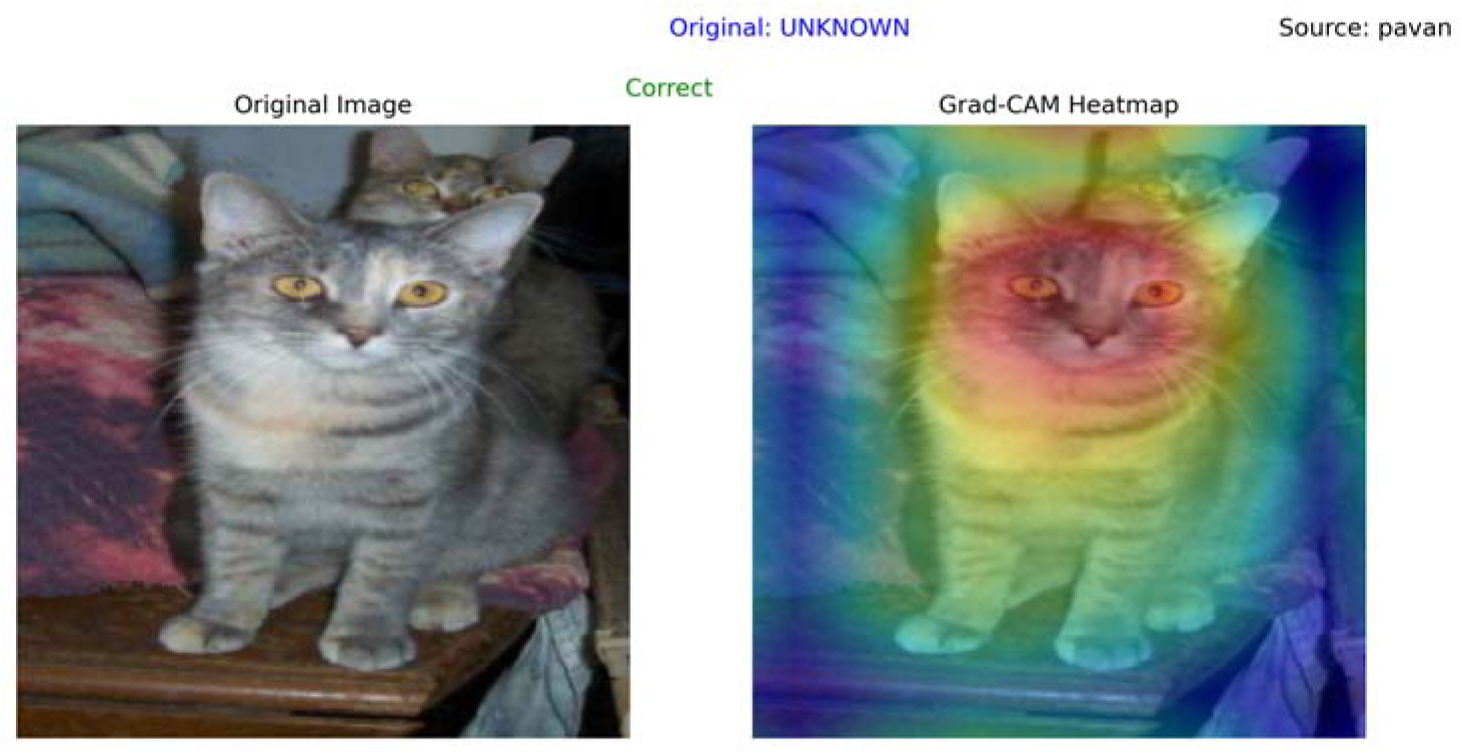
Correct Unknown prediction

#### DenseNet-121 Grad-CAM Analysis

To analyze the model’s predictions, 7 visualizations were created:

- **4 Correct Predictions**: One from each class (Normal: Figure 11, Pneumonia: Figure 12, Tuberculosis: Figure 15, Non-X-ray: Figure 16).
- **3 Incorrect Predictions**: One misclassified example per class (Normal: Figure 10, Pneumonia: Figure 13, Tuberculosis: Figure 14). No misclassifications occurred in the Non-X-ray class

**Figure 10.**
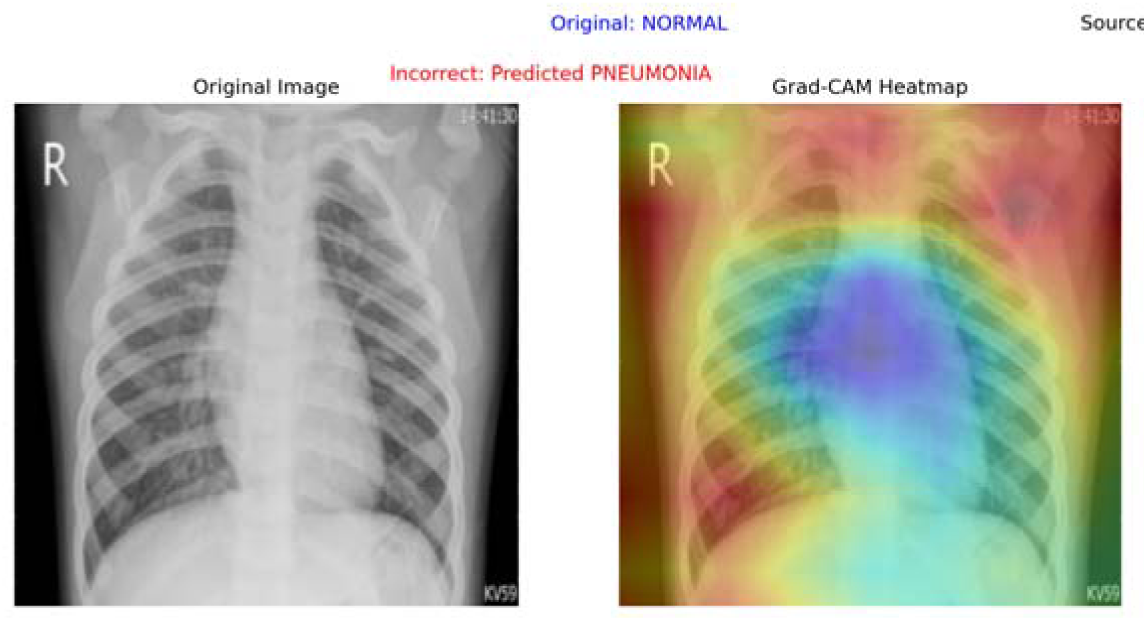
Incorrect Pneumonia Prediction of Normal

**Figure 11.**
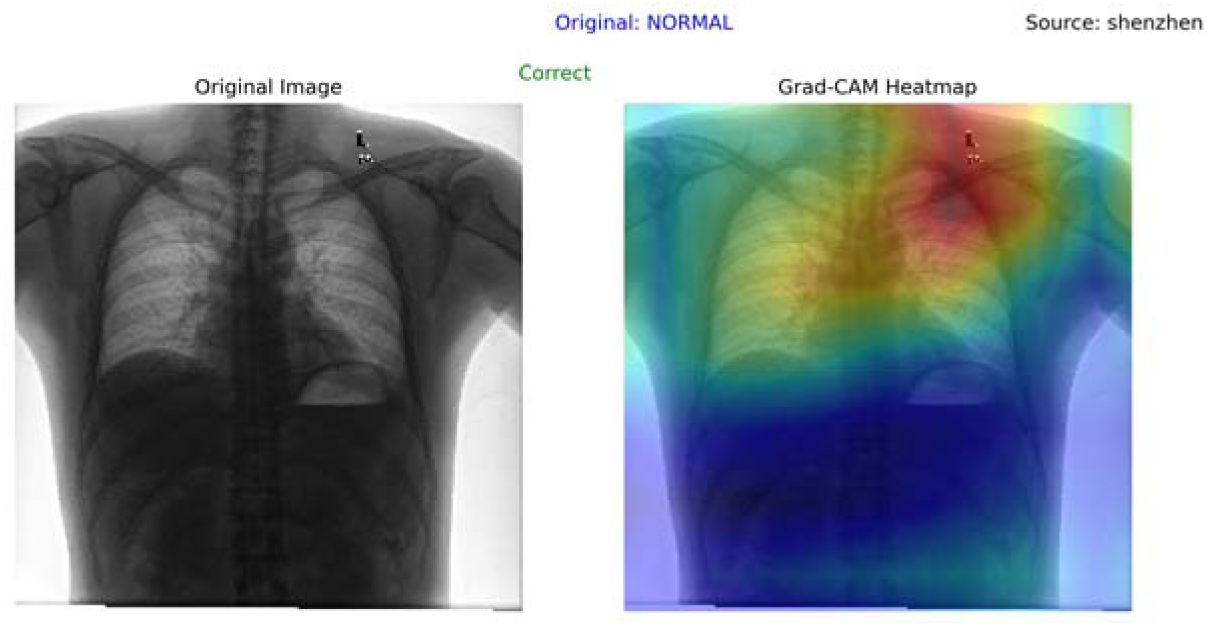
Correct Normal Predicted

**Figure 12.**
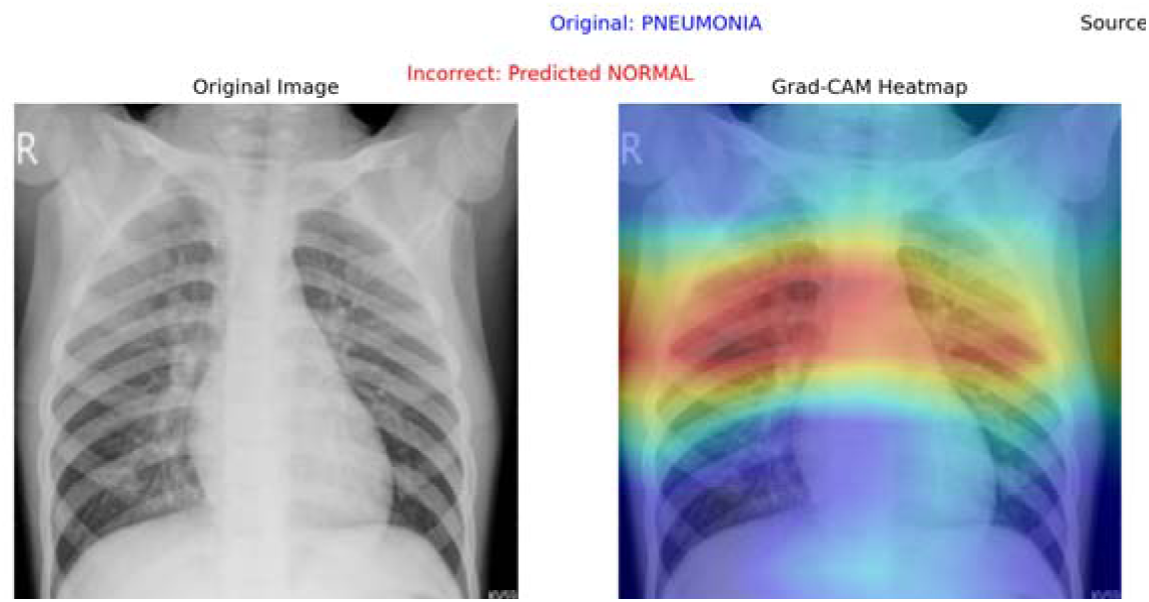
Incorrect Normal Prediction of Pneumonia

**Figure 13.**
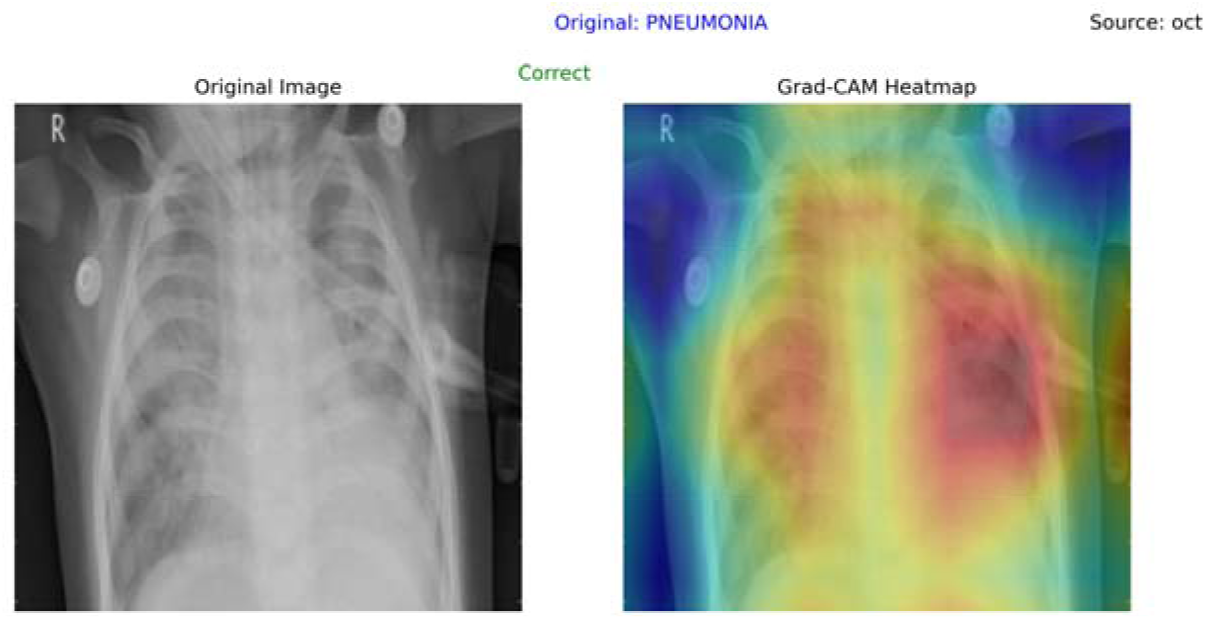
Correct Pneumonia Predicted

**Figure 14.**
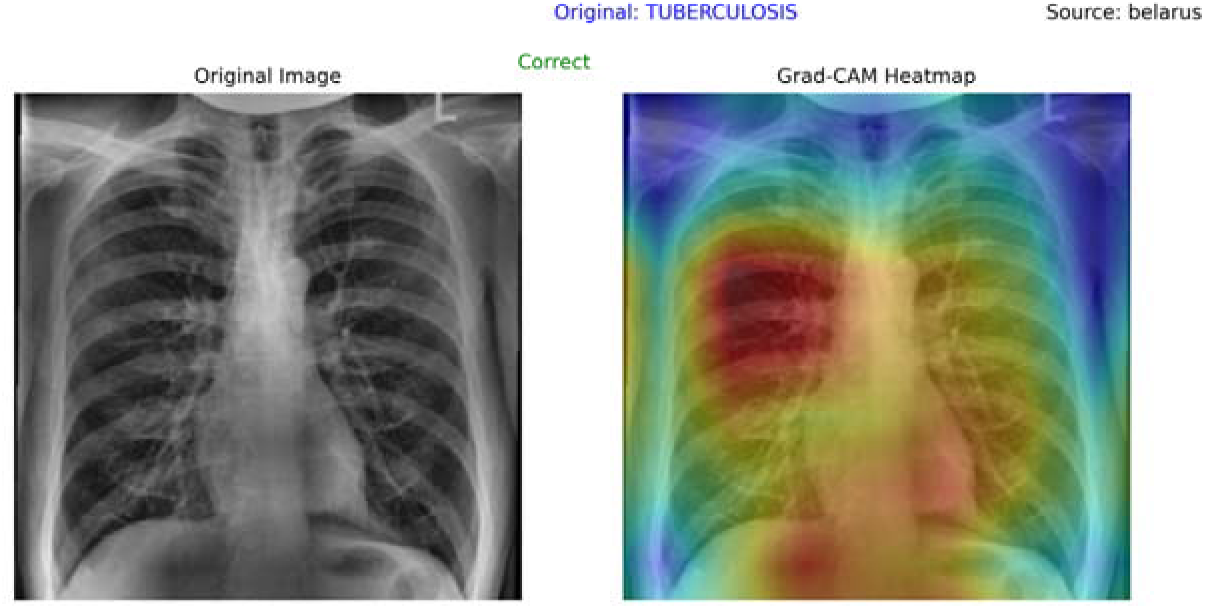
Correct Normal Tuberculosis

**Figure 15.**
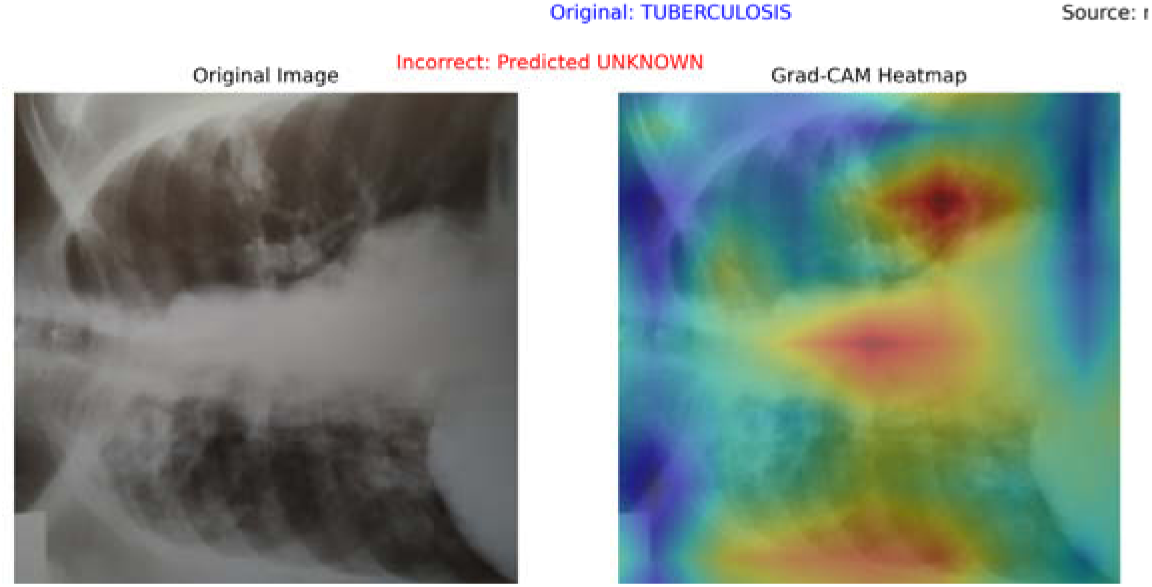
Incorrect Unknown Prediction of Tuber ulosis

**Figure 16.**
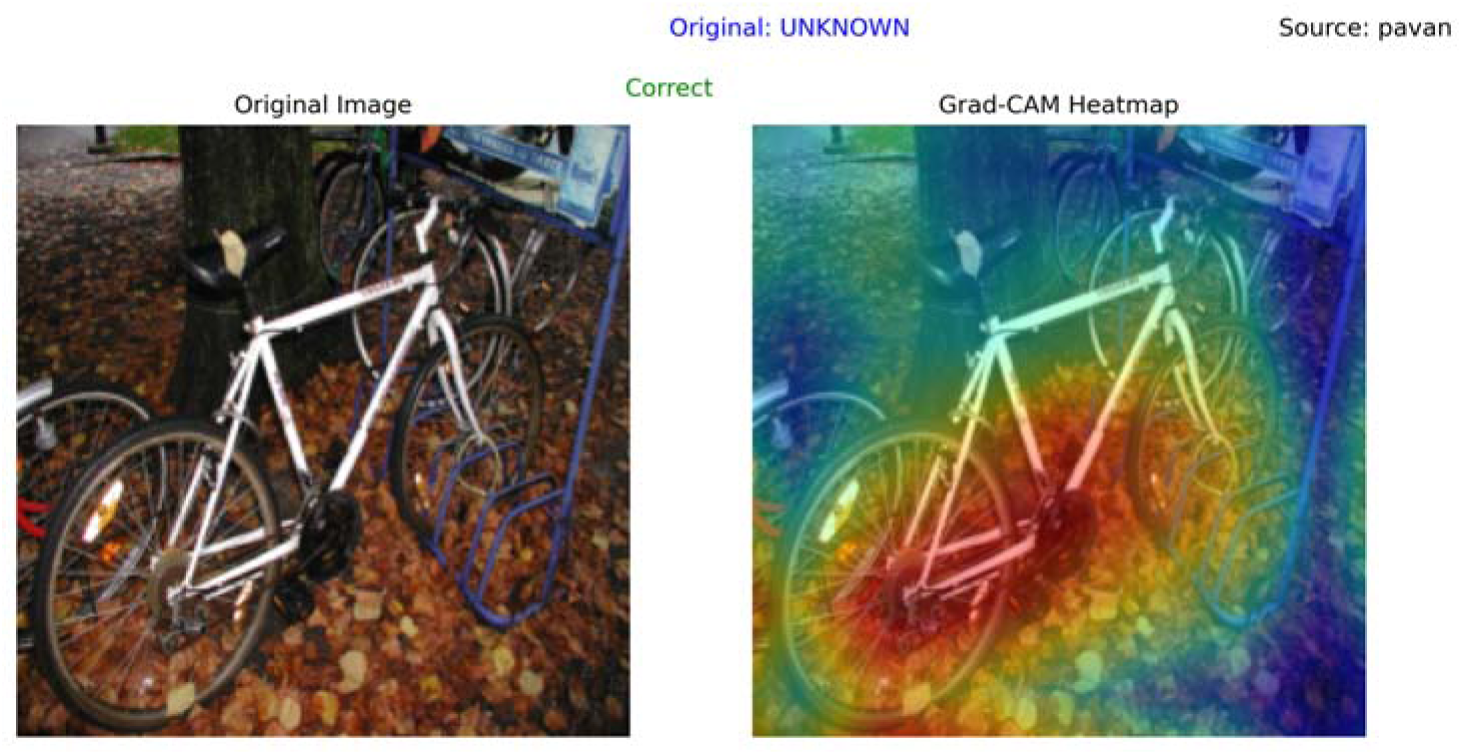
Correct Unknown Predicted

#### Key Insights

- **Non-X-ray Class**: The “Non-X-ray” class was expected to be relatively easy to classify, given its clear distinction. However, DenseNet-121 misclassified a sample as “non-X-ray” in Figure 14, which represents an image with a vertical chest X-ray. The model mistakenly classified this as “Non-X-ray,” even though it was a valid X-ray image, highlighting a potential weakness in generalization. A “non-X ray” classification might have been expected, which raises questions about the model’s ability to distinguish between X-ray and non-X-ray images. In contrast, Figure 17 shows that ResNet-18 was able to classify the same vertical chest X-ray correctly, demonstrating its superior handling of image orientation and positioning.
- Vertical vs. Horizontal Chest X-rays: The misclassification in DenseNet-121 with the vertical X-ray raises concerns about the model’s robustness to different image orientations. One key concern is that the dataset consists primarily of horizontal X-ray images, which could explain why DenseNet-121 struggles with vertical images. ResNet-18, however, appeared more adaptable to such variations, correctly identifying the class despite the orientation in Figure 17. This highlights ResNet-18’s superior generalization and robustness, making it more reliable for real-world clinical applications, where images might not always follow standardized orientations and could potentially include varying orientation.

**Figure 17.**
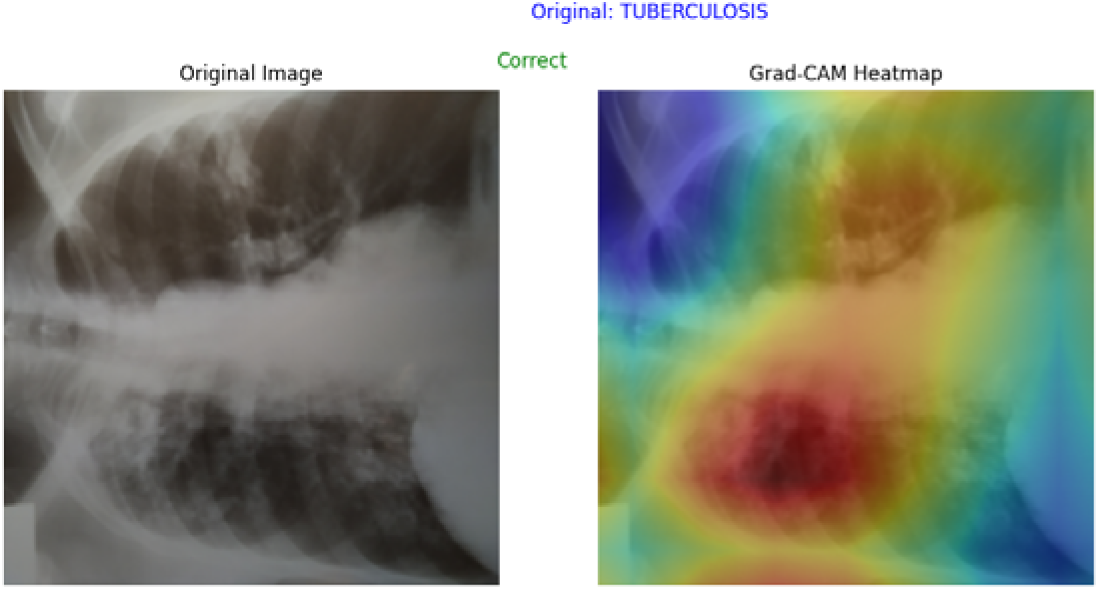
RseNet18 Identifying the same image correctly

### 5.5) Supplementary Figures

This section provides graphical representations of the training and evaluation processes for both ResNet-18 and DenseNet-121 models. The figures offer insights into the models’ performance during training, validation, and testing phases

**Figure 18.**
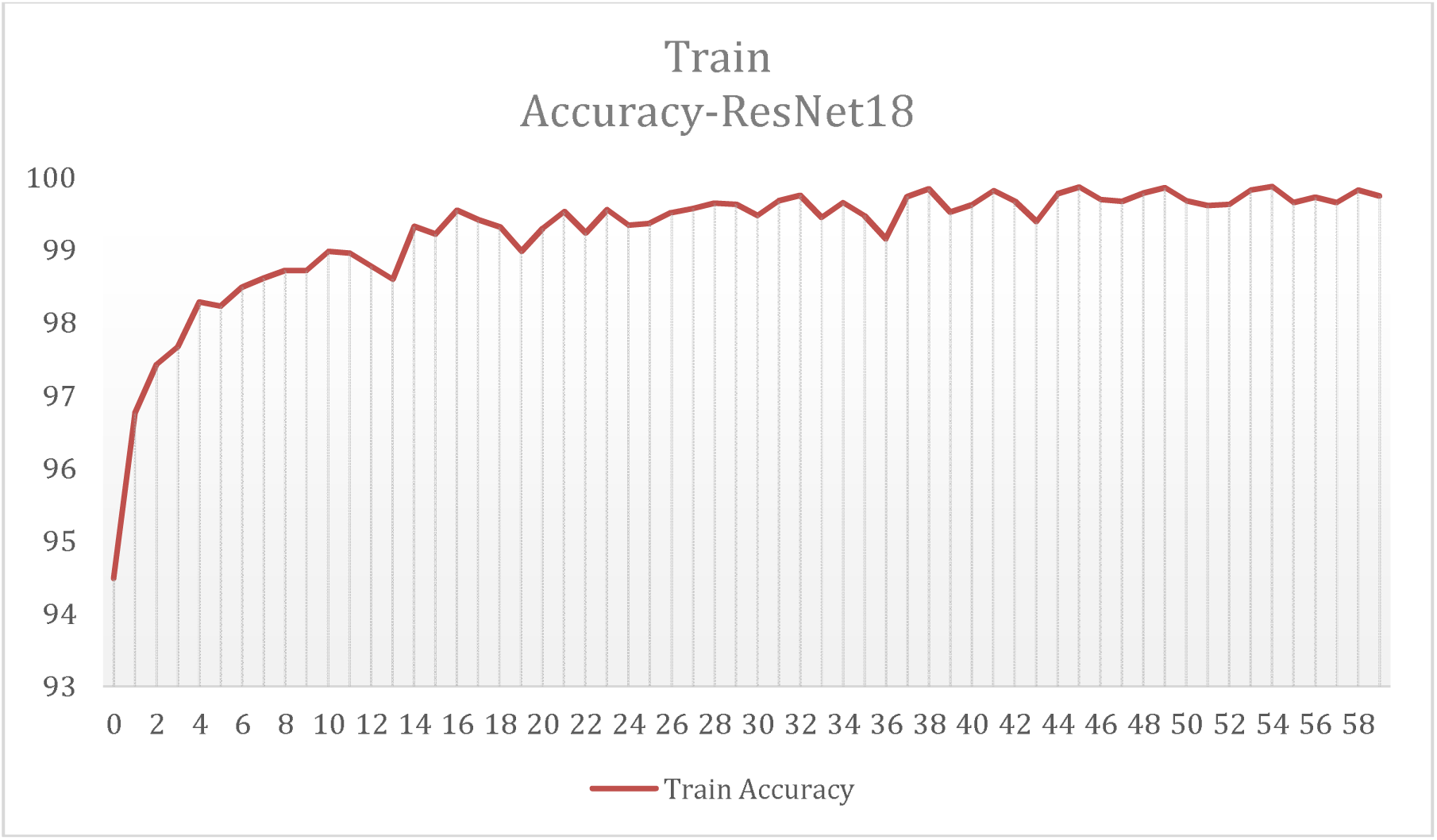
Training Accuracy ResNet18

**Figure 19.**
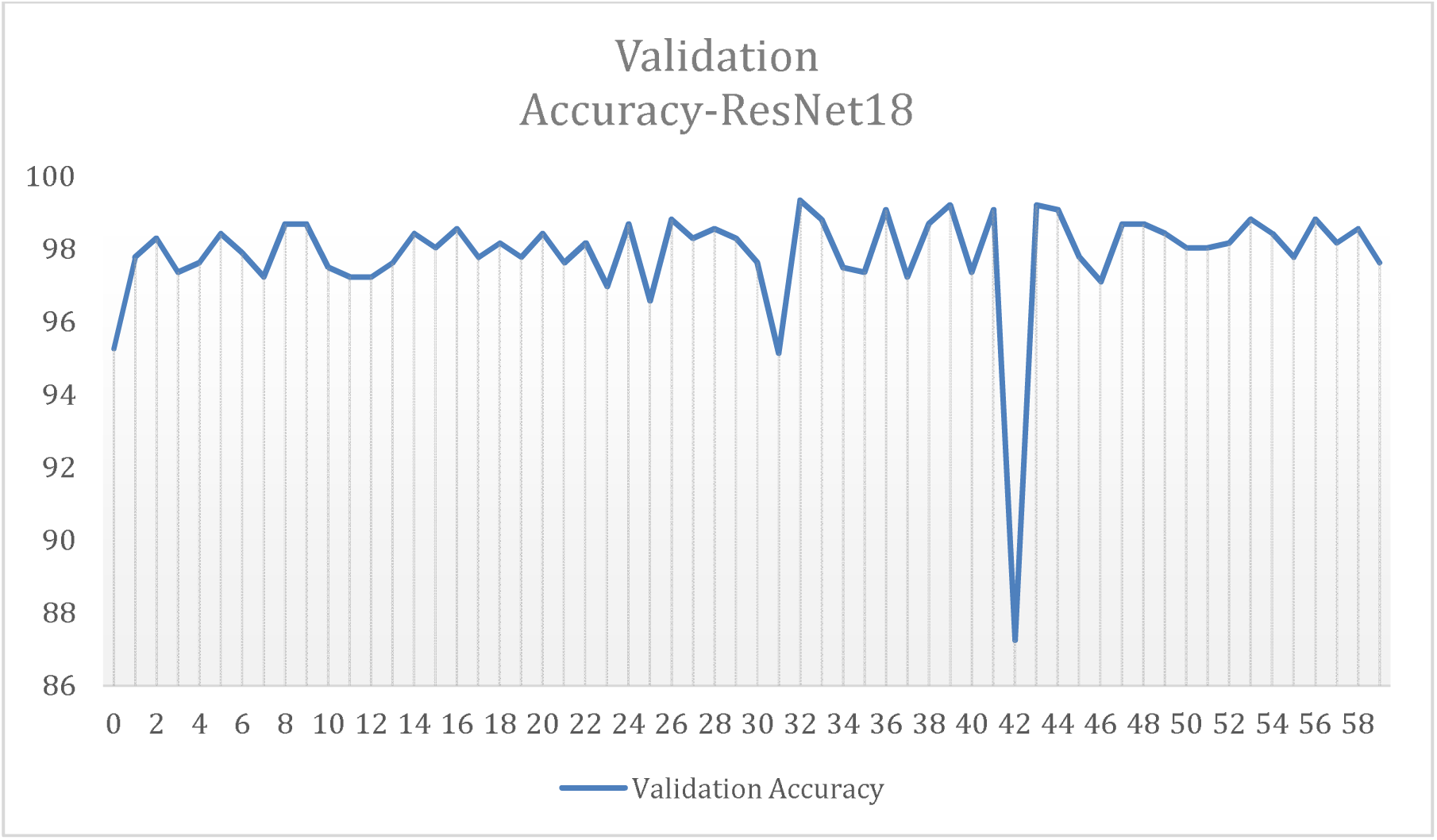
Validation Accuracy ResNet18

#### Training Metrics

- Graphs depicting **training accuracy** and **training loss** over the course of all epochs are presented. These graphs demonstrate how each model progressively improved its ability to classify the data and reduced the loss as training progressed.

**Figure 20.**
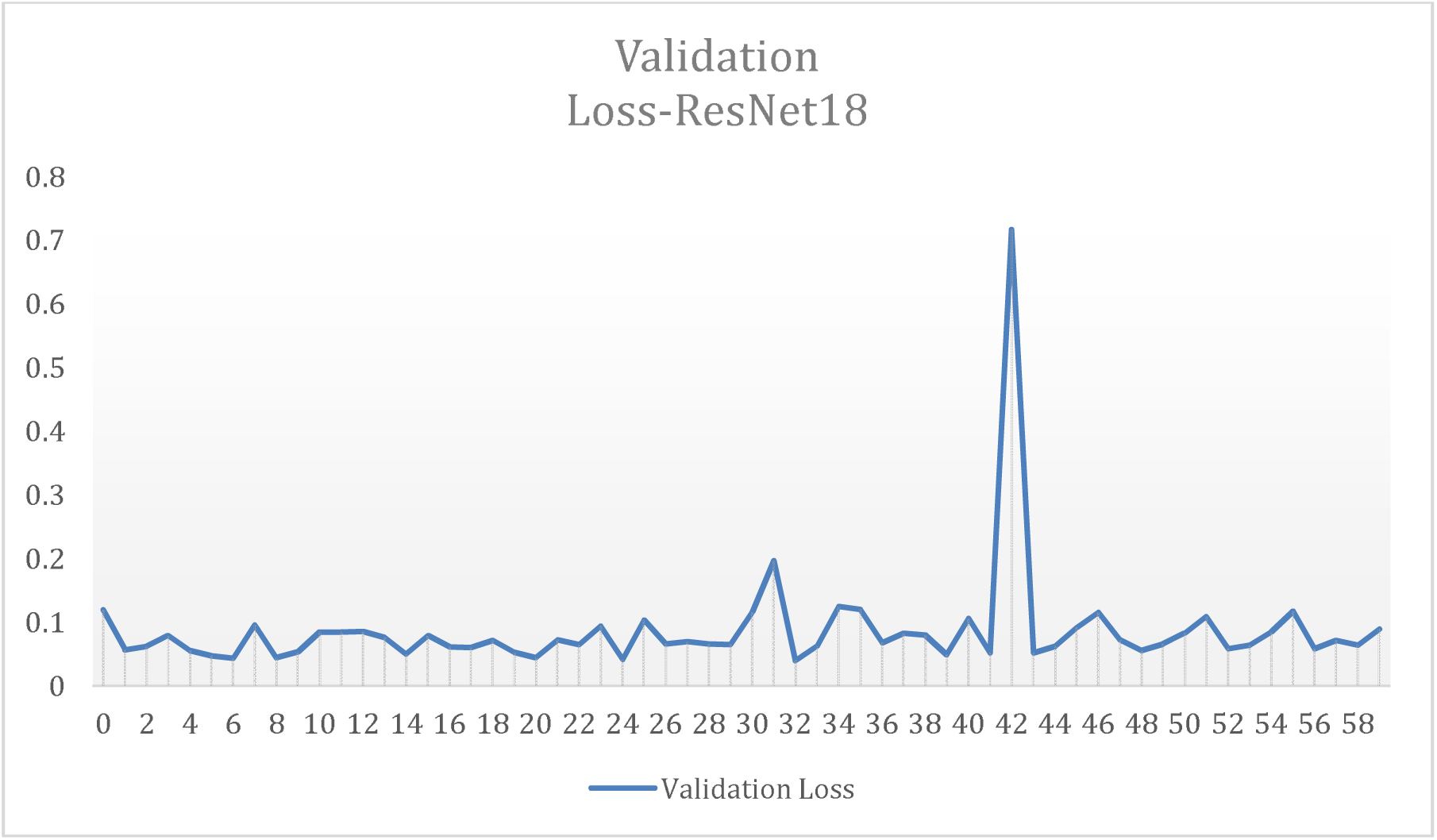
Validation Loss ResNet18

**Figure 21.**
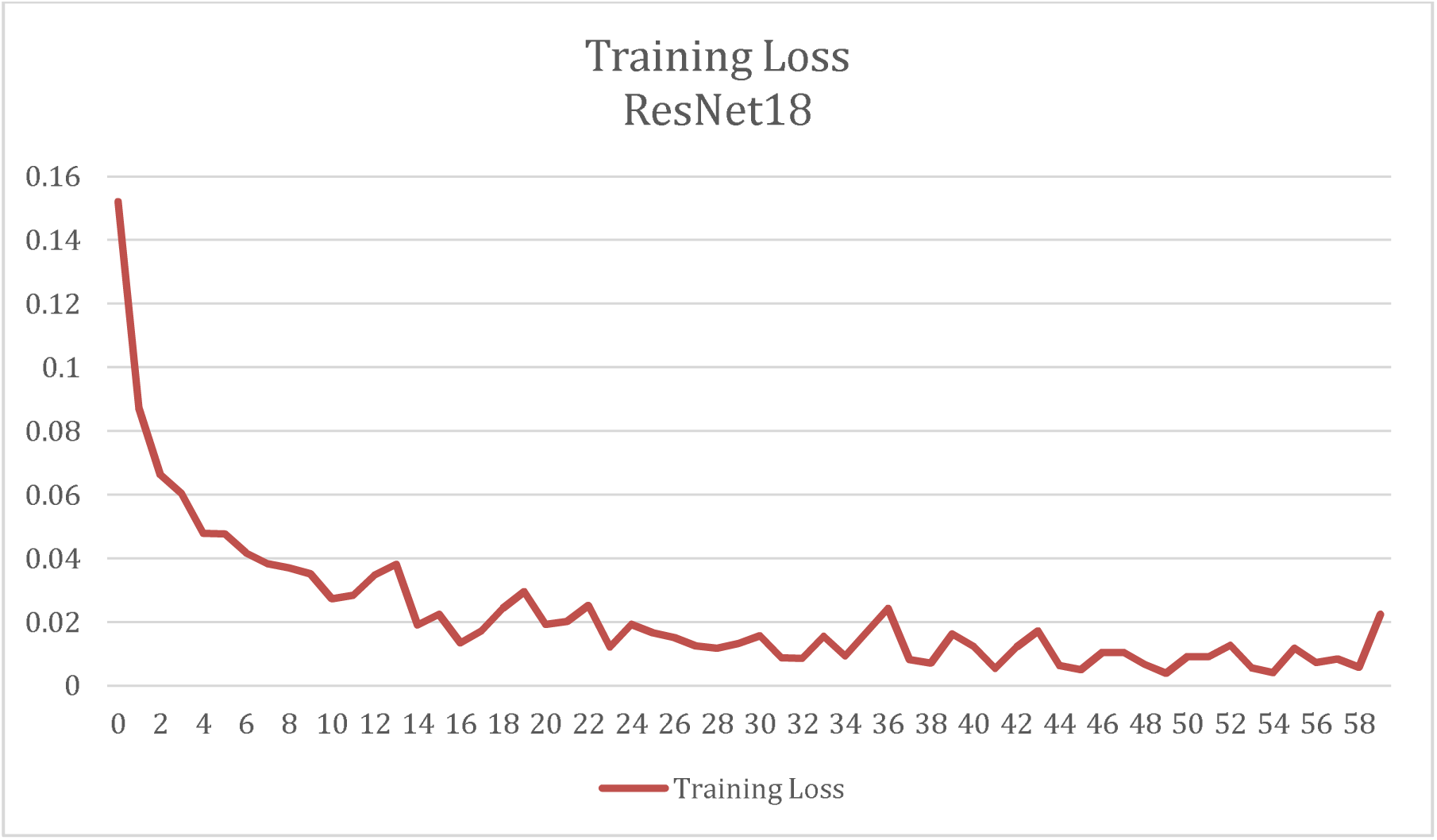
Training Loss ResNet18

#### Validation Metrics

- Figures illustrating the validation accuracy and validation loss for both models over all epochs. These plots provide a clear view of the models’ generalization capabilities and highlight their performance on unseen validation data during training.

**Figure 22.**
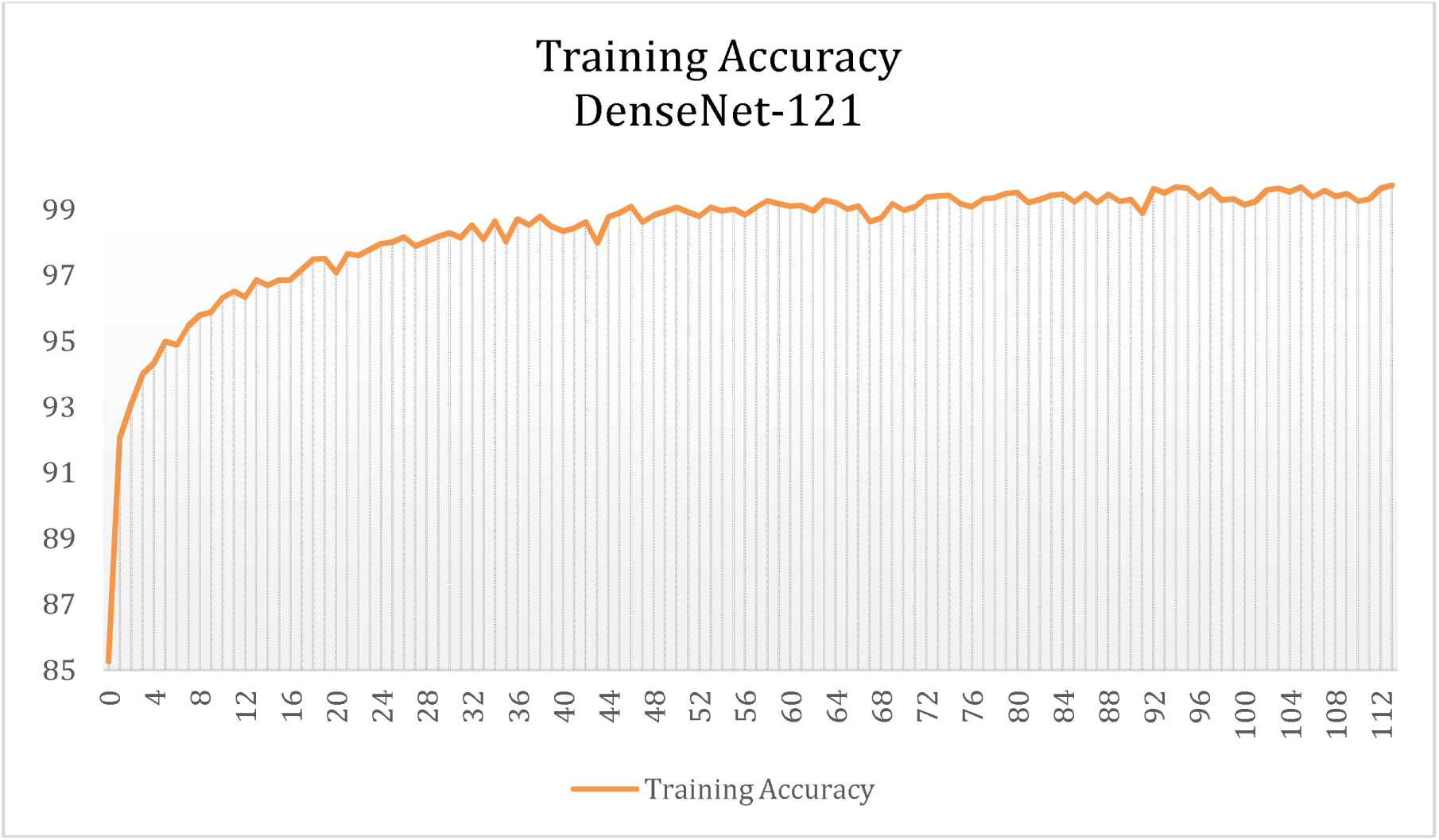
Training Accuracy DenseNet-121

**Figure 23.**
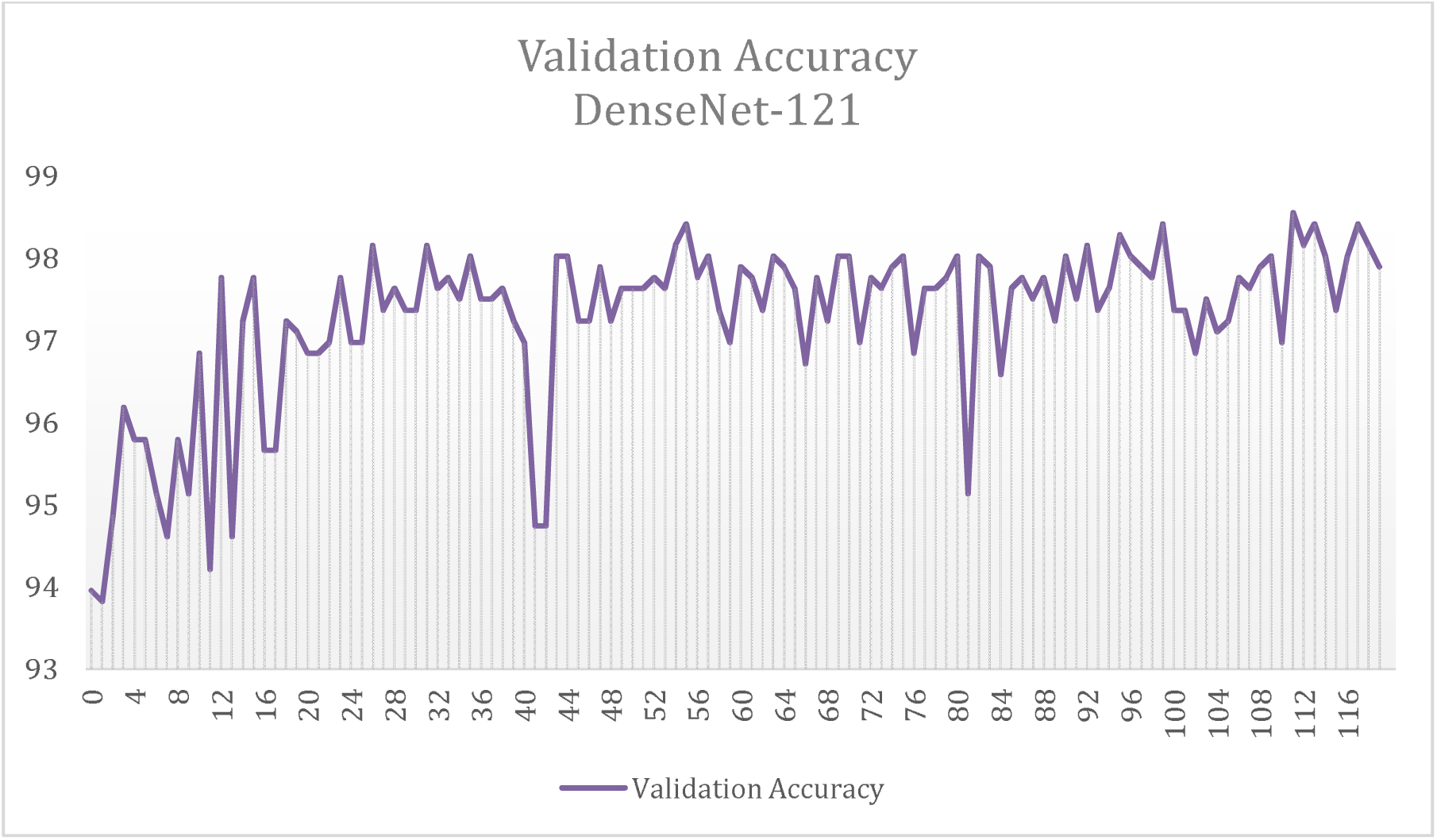
Validation Accuracy DenseNet-121

#### Testing Results

- Final evaluation results are visualized using graphs of the **test accuracy** and **test loss** for each model. These figures showcase how well ResNet-18 and DenseNet-121 performed on completely unseen data, reflecting their robustness and classification effectiveness.

**Figure 24.**
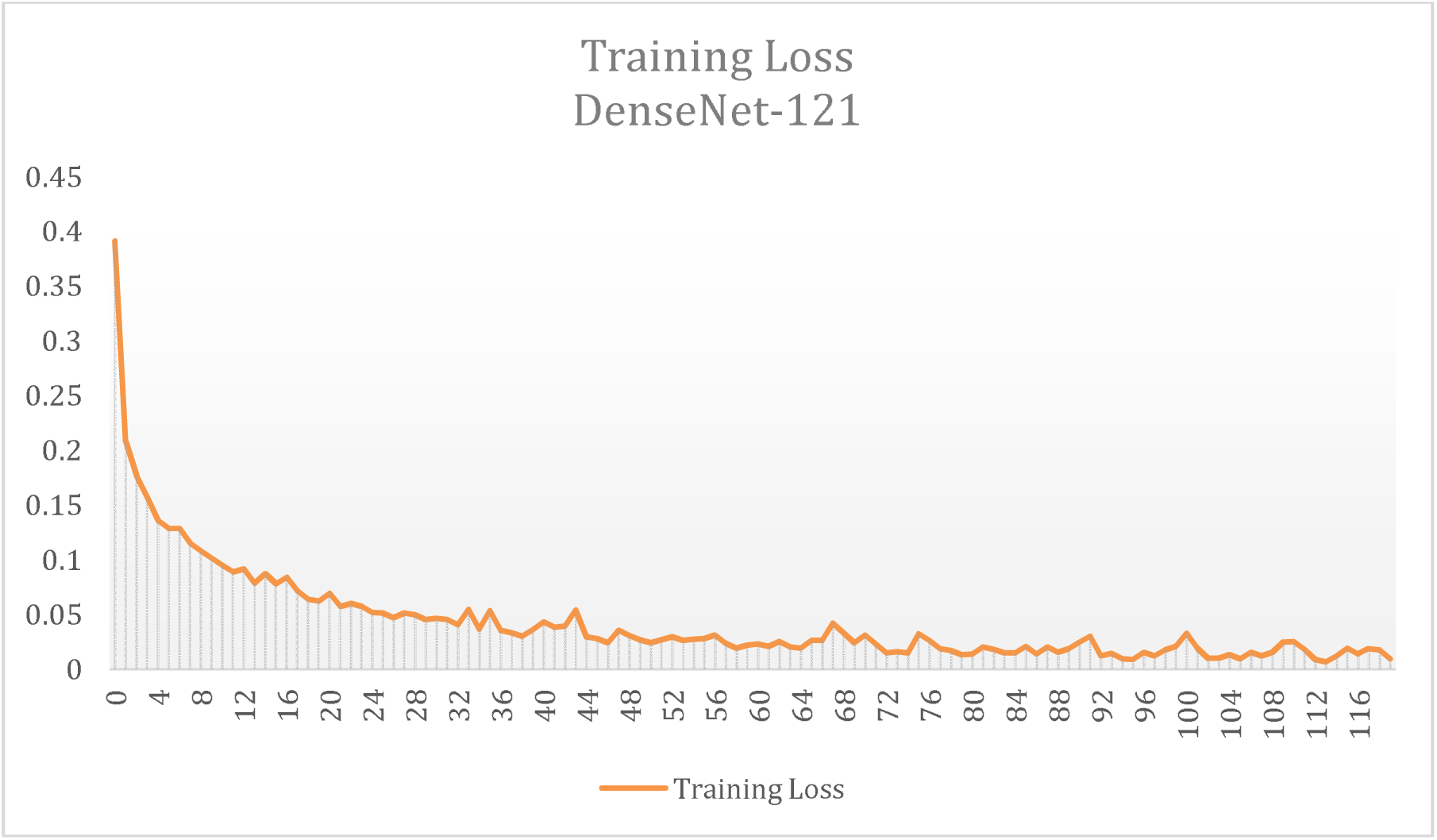
Training Loss DenseNet-121

**Figure 25.**
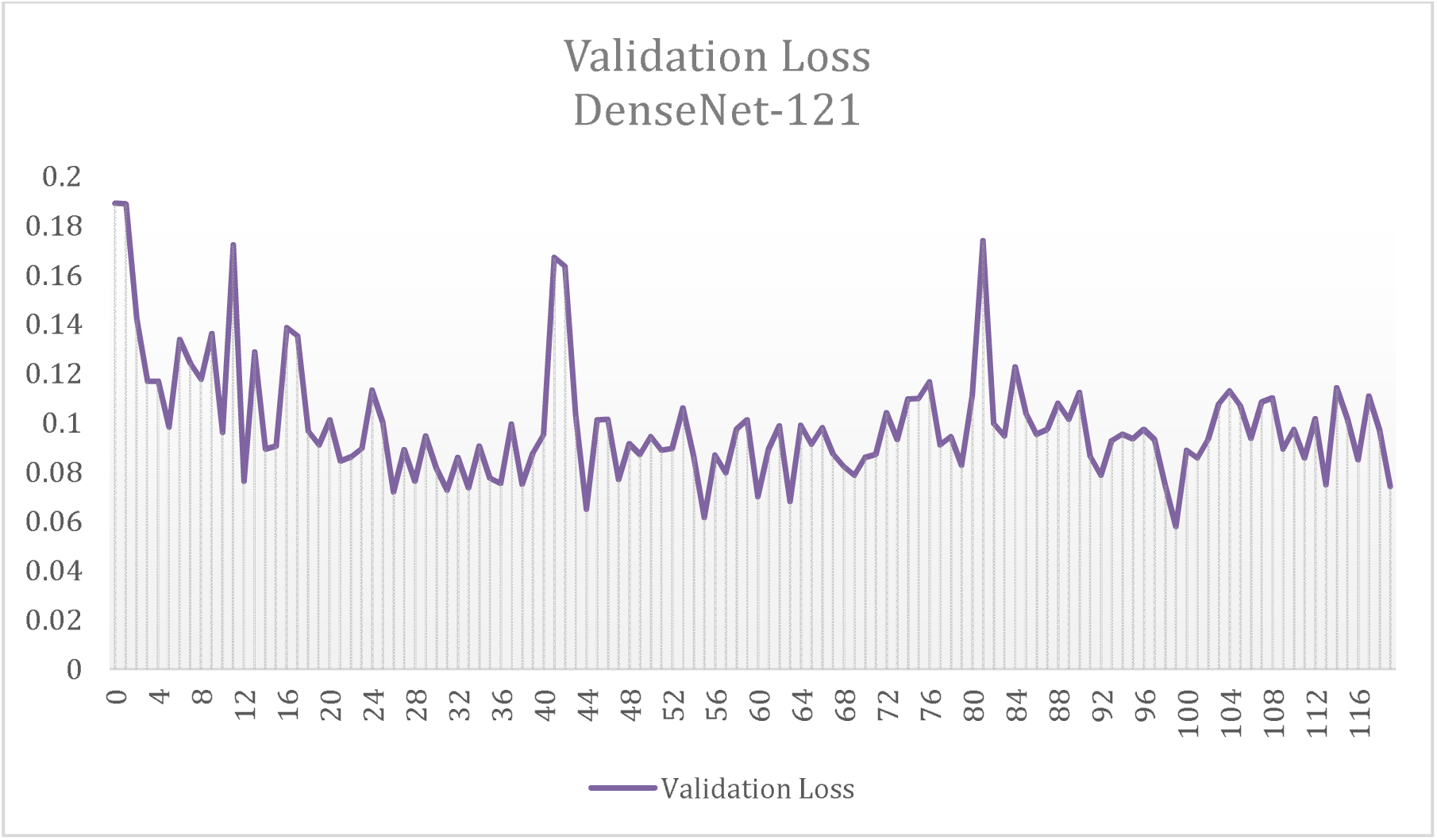
Validation Loss DenseNet-121

**Figure 26.**
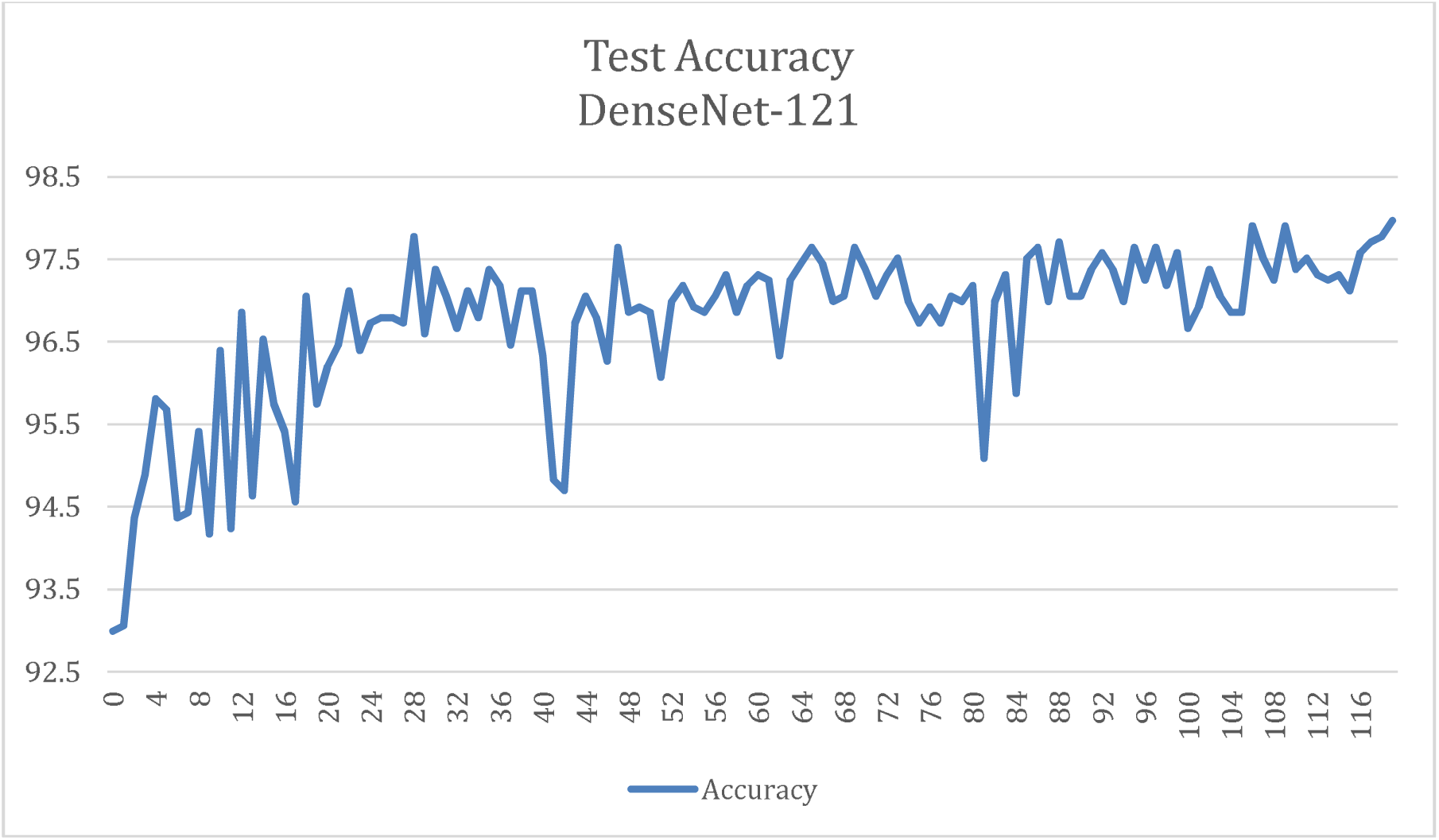
Test Accuracy DenseNet-121

**Figure 27.**
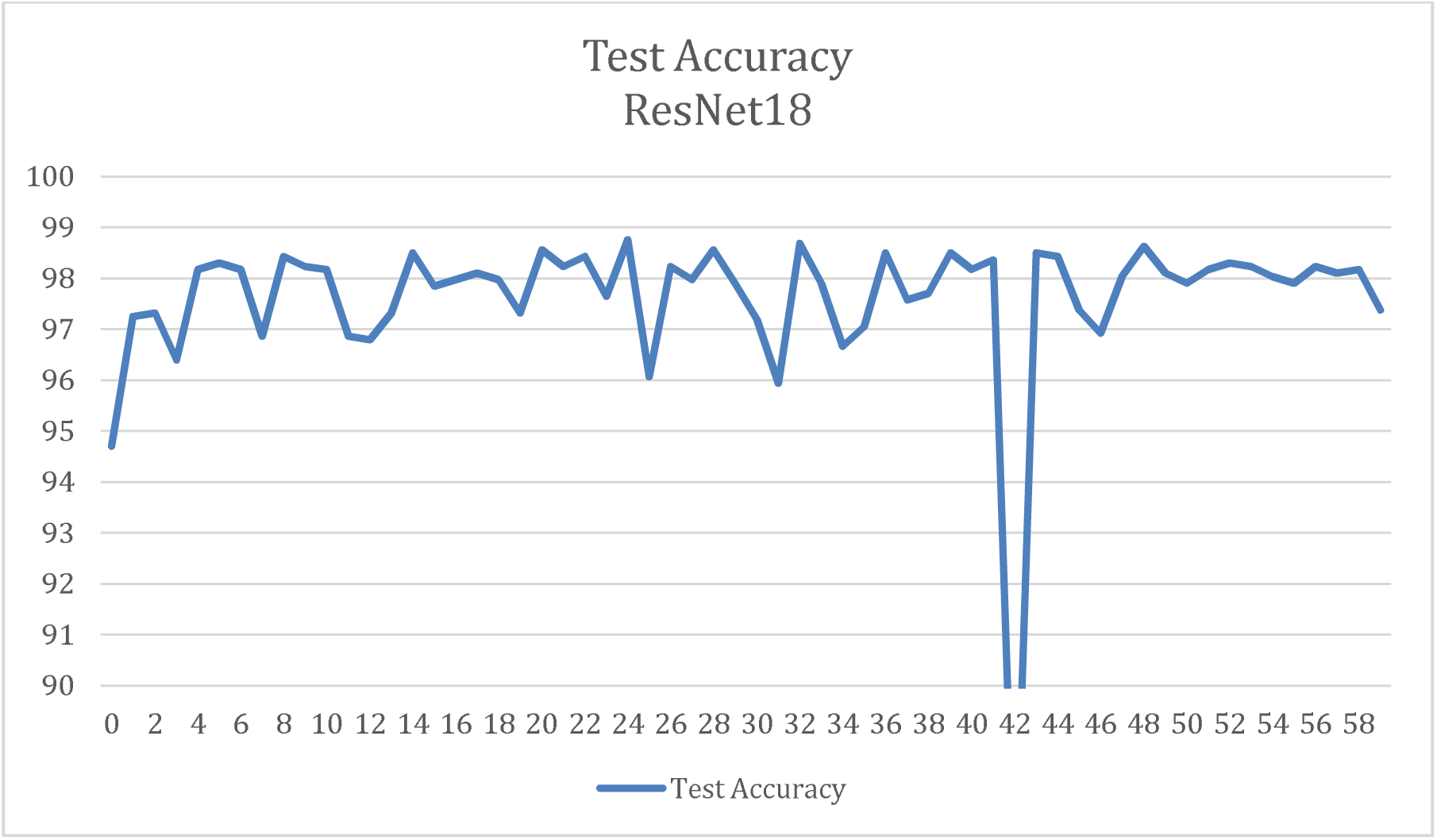
Test Accuracy ResNet18

### 5.5) Time and Computational Analysis

This section presents a comparative analysis of the time and computational efficiency of ResNet-18 and DenseNet-121, focusing on their performance during inference and computational resource utilization.

- Inference Time: ResNet-18 demonstrates superior inference speed, processing a single image in **0.24 seconds**, compared to **0.31 seconds** for DenseNet-121. This makes ResNet-18 approximately **20–25% faster**, which is critical for real-time or resource-constrained environments.
- Computational Complexity:

- Parameters: ResNet-18 has 11.7 million trainable parameters, slightly more than DenseNet-121, which has 8.0 million parameters.
- FLOPs (Floating Point Operations): ResNet-18 requires 1.8 billion FLOPs, compared to 2.8 billion FLOPs for DenseNet-121, indicating that ResNet-18 has lower computational demands.
- Memory Usage: DenseNet-121 consumes 30.8 MB of memory, slightly less than ResNet-18, which uses 45 MB. However, both models are significantly lighter than larger architectures such as DenseNet-201 (78 MB) and ResNet-152 (230 MB).
- Performance vs. Complexity: ResNet-18 strikes an excellent balance between computational efficiency and performance. It achieves high accuracy with lower computational requirements, making it highly suitable for deployment in resource-constrained environments. When deployed in a containerized Docker server and run solely on CPU, ResNet-18 achieves an inference time as low as **0.02 seconds per image**, making it highly efficient for local hardware setups while maintaining medical data integrity and ensuring compliance with security standards.
- Model Selection Recommendations:

- **Time-critical applications:** ResNet-18 is recommended due to its faster inference speed.
- **Resource-constrained applications:** DenseNet-121 may be preferred for memory-limited scenarios, though ResNet-18 is still suitable for computational efficiency.
- **Performance-driven applications:** For tasks demanding higher accuracy, ResNet-18 is the better choice.

### 5.6) Direct Comparison with State-of-the-Art Models

This section presents a performance comparison between our proposed ResNet-18 model and various state-of-the-art methods for detecting pneumonia and tuberculosis from chest X-rays. The analysis evaluates key metrics—accuracy, precision, recall, F1-score—and computational efficiency, highlighting ResNet-18’s advantages for real-world clinical applications.

A. Key Results of ResNet-18:

1. **Overall Accuracy:** Achieved 98.76%, surpassing complex models like UNet+DenseNet201 [25] (98.60%) and Vision Transformers (97.61%) [9], while avoiding segmentation overhead.
2. Class-Specific Metrics:
3. *Normal:* Precision 98.53%, Recall 98.00%, F1-Score 98.26%.
4. *Pneumonia:* Precision 97.91%, Recall 98.83%, F1-Score 98.37%.
5. *Tuberculosis:* Precision 99.52%, Recall 99.28%, F1-Score 99.40%.
6. *Non-X-ray:* Achieved perfect scores with Precision, Recall, and F1-Score all at 100%.
7. **Efficiency:** Processes images 20% −25% faster than DenseNet-121 and achieves the lowest trainable parameters in the study, enabling deployment in resource-constrained environments. With a computational cost of 1.8B FLOPs, it maintains practical usability while producing 97.91% overall accuracy across four classes (see Section 5.3 for detailed reports).

**Table 3.**
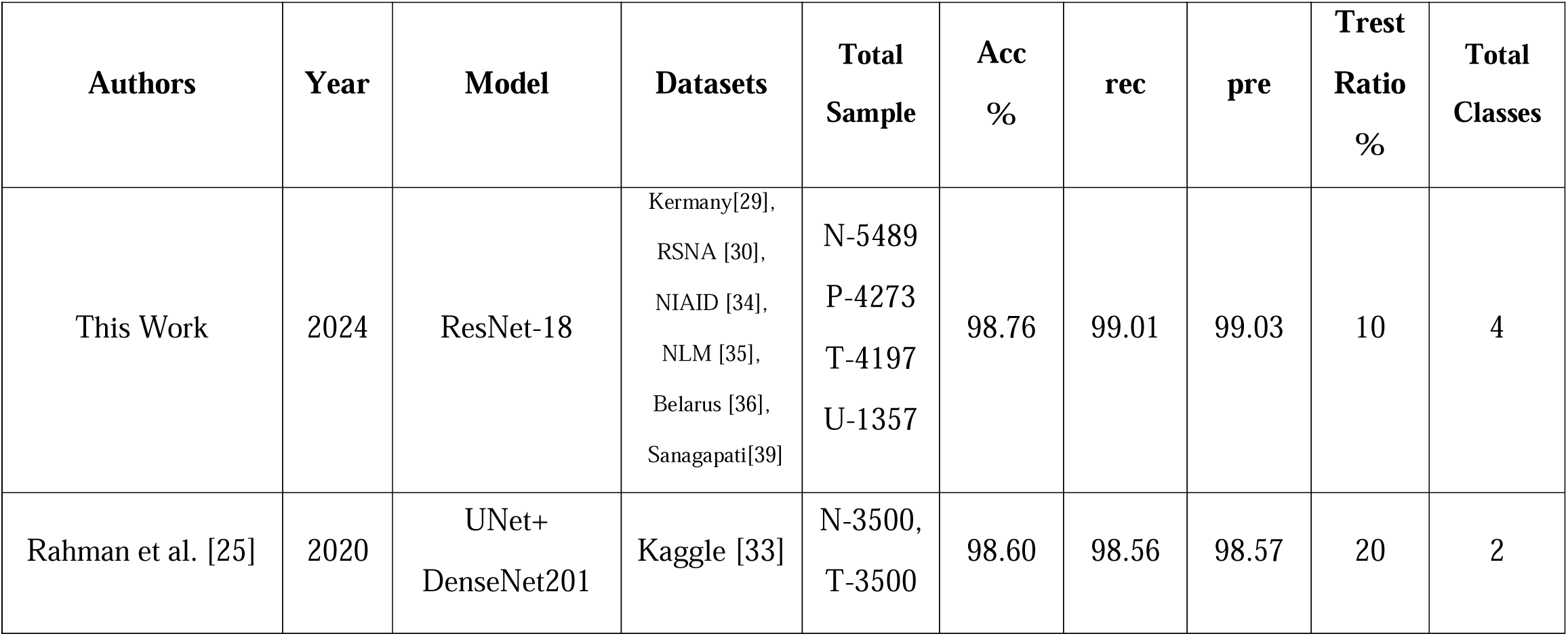
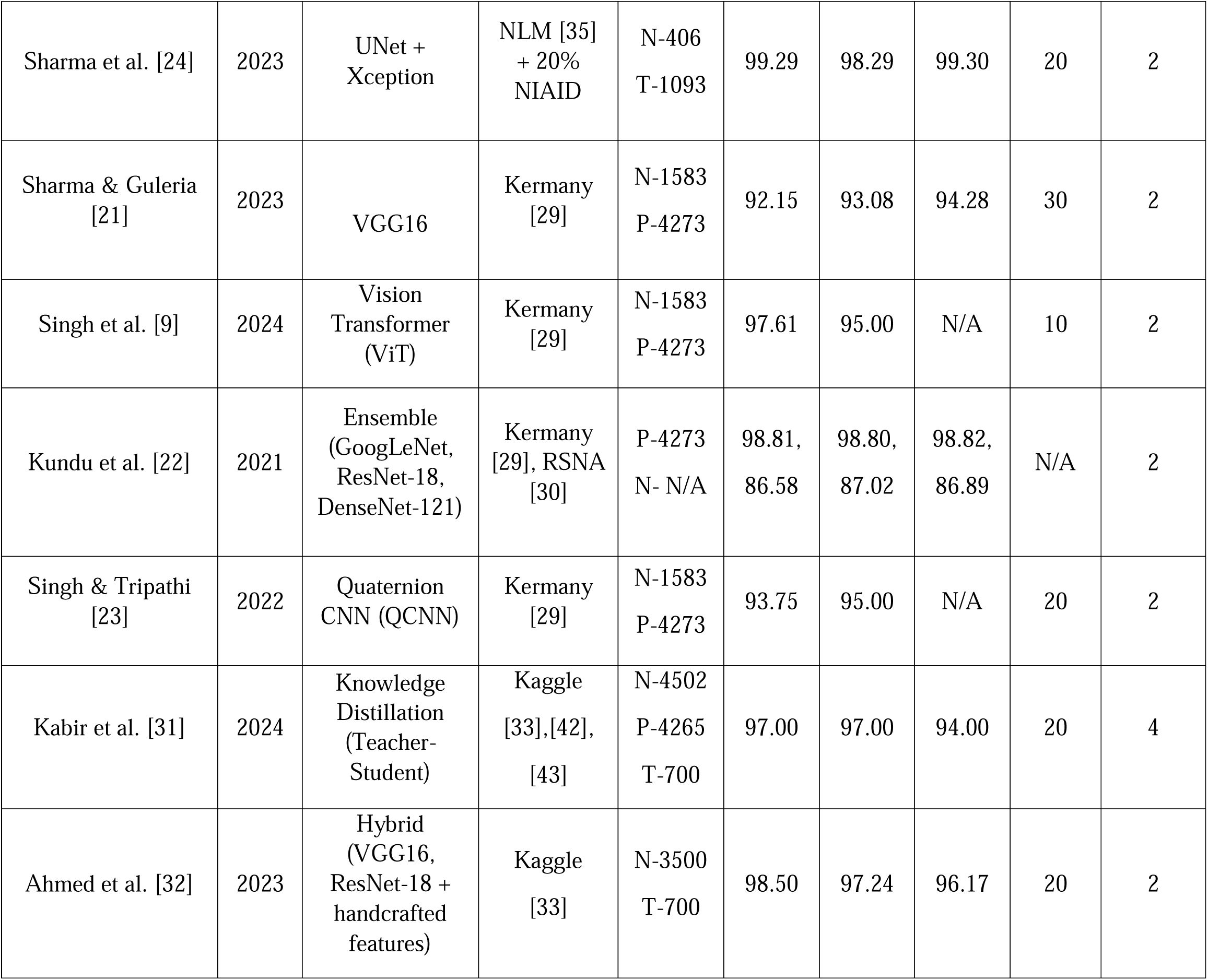
Comparison With State-of-the-art.

| Authors | Year | Model | Datasets | Total Sample | Acc % | rec | pre | Trest Ratio % | Total Classes |
| --- | --- | --- | --- | --- | --- | --- | --- | --- | --- |
| This Work | 2024 | ResNet-18 | Kermany[29],<br>RSNA [30],<br>NIAID [34],<br>NLM [35],<br>Belarus [36],<br>Sanagapati[39] | N-5489<br>P-4273<br>T-4197<br>U-1357 | 98.76 | 99.01 | 99.03 | 10 | 4 |
| Rahman et al. [25] | 2020 | UNet+<br>DenseNet201 | Kaggle [33] | N-3500,<br>T-3500 | 98.60 | 98.56 | 98.57 | 20 | 2 |
| Sharma et al. [24] | 2023 | UNet + Xception | NLM [35] + 20% NIAID | N-406<br>T-1093 | 99.29 | 98.29 | 99.30 | 20 | 2 |
| Sharma & Guleria [21] | 2023 | VGG16 | Kermany [29] | N-1583<br>P-4273 | 92.15 | 93.08 | 94.28 | 30 | 2 |
| Singh et al. [9] | 2024 | Vision Transformer (ViT) | Kermany [29] | N-1583<br>P-4273 | 97.61 | 95.00 | N/A | 10 | 2 |
| Kundu et al. [22] | 2021 | Ensemble (GoogLeNet, ResNet-18, DenseNet-121) | Kermany [29], RSNA [30] | P-4273<br>N- N/A | 98.81,<br>86.58 | 98.80,<br>87.02 | 98.82,<br>86.89 | N/A | 2 |
| Singh & Tripathi [23] | 2022 | Quaternion CNN (QCNN) | Kermany [29] | N-1583<br>P-4273 | 93.75 | 95.00 | N/A | 20 | 2 |
| Kabir et al. [31] | 2024 | Knowledge Distillation (Teacher-Student) | Kaggle [33],[42], [43] | N-4502<br>P-4265<br>T-700 | 97.00 | 97.00 | 94.00 | 20 | 4 |
| Ahmed et al. [32] | 2023 | Hybrid (VGG16, ResNet-18 + handcrafted features) | Kaggle [33] | N-3500<br>T-700 | 98.50 | 97.24 | 96.17 | 20 | 2 |

**B. Performance Comparison:** ResNet-18 consistently outperforms state-of-the-art models in multiclass tasks:

1. It achieves higher accuracy (98.76%) compared to DenseNet201-based models (98.60%) [25], Vision Transformers (97.61%) [9], and hybrid approaches [32], which combine CNNs with handcrafted features.
2. By maintaining a simple architecture, ResNet-18 avoids the complexities of hybrid or ensemble models without compromising robustness.

**C. Generalizability:** ResNet-18 demonstrates strong adaptability due to training on a diverse dataset that includes Kermany, RSNA, NIAID, NLM, Belarus, and non-X-ray data. Its ability to classify an “Non-X-ray” class further underscores its practicality for real-world clinical scenarios.

**D.** Comparison Highlights:

1. **Accuracy and Robustness:** Achieves the highest accuracy (98.76%) among multiclass classifiers, with balanced performance across all categories.
2. **Computational Efficiency:** Processes images faster than any other study, combining high accuracy with low computational demands for practical deployment.
3. **Adaptability:** Delivers consistent results across diverse data sources and imaging conditions, outperforming methods reliant on smaller or less varied datasets.

### 5.7) Deployment Strategy

ResNet-18 can be deployed with trained weights in multiple ways. One option is to convert the model weights to the Open Neural Network Exchange (ONNX) format, allowing it to run locally on any resource-constrained machine with ease. This approach can be combined with programming languages such as C++, C#, or JavaScript.

The strategy I employed in for my application was to combine the model and weights, wrap them with an asynchronous Tornado server, and then compile the entire setup into a Docker container. This container includes all necessary dependencies, ensuring that the deployment works seamlessly across different machines, regardless of the environment.

## Section 6: Conclusion and Future Work

This study demonstrates the effectiveness of ResNet-18 and DenseNet-121 models for the multiclass classification of chest X-rays, specifically targeting Normal, Pneumonia, Tuberculosis, and Non-X-ray categories. By incorporating diverse datasets and exploring the impact of class weights, the research provides valuable insights into the trade-offs between accuracy, computational efficiency, and generalization.

### 6.1) Key Findings

1. **Multiclass Classification:** The multiclass classification approach significantly outperformed binary classification, achieving better predictive accuracy by considering multiple disease categories. This approach highlights the importance of expanding beyond binary models for more comprehensive diagnostic tasks, offering broader applicability in clinical settings.
2. **Handling non-X-ray Anomalies:** The inclusion of “Non-X-ray” anomalies, such as non-X-ray images, improved the models’ ability to generalize across diverse data types. This adaptation proved critical in distinguishing valid medical images from irrelevant or erroneous inputs, enhancing robustness and real-world applicability, especially in clinical environments where data quality may vary.
3. **ResNet-18 Performance:** ResNet-18 consistently outperformed DenseNet-121, Vision Transformers (ViT), VGG16, DenseNet-201, and ensemble methods, achieving the highest test accuracy of 98.76%. Its relatively simple architecture delivered high performance while maintaining computational efficiency. Furthermore, ResNet-18 demonstrated robustness to diverse datasets and imaging conditions, making it a strong candidate for deployment.
4. **DenseNet-121 as an Alternative:** While DenseNet-121 showed slightly lower accuracy compared to ResNet-18, it remained competitive, particularly for applications where memory efficiency is a priority. However, its slower inference times and higher computational complexity suggest that ResNet-18 is better suited for real-time applications and resource-constrained environments.
5. **Efficiency and Practicality:** ResNet-18 exhibited superior inference speed, processing images 20–25% faster than DenseNet-121, with a significantly lower computational cost (1.8 billion FLOPs). This makes ResNet-18 particularly suitable for resource-constrained environments, such as real-time clinical applications.
6. **Clinical Relevance:** Grad-CAM visualizations confirmed the models’ ability to focus on clinically relevant regions of input images, ensuring interpretability and reliability for real-world applications. Additionally, the classification of the “Non-X-ray” category demonstrated the models’ adaptability to diverse imaging sources, making them robust for clinical deployment.
7. **Impact of Weighted Loss Function:** The inclusion of a weighted loss function did not significantly improve performance and, in some cases, led to slight overfitting, particularly with ResNet-18. Models trained without class weights exhibited better generalization, suggesting that dataset balance may have mitigated the need for reweighting.
8. **Class-Specific Insights:** Both models excelled at classifying the “Non-X-ray” class with 100% precision and recall, while slight misclassifications occurred with “Normal” and “Pneumonia.” ResNet-18 also demonstrated superior handling of diverse image orientations, which DenseNet-121 struggled with, highlighting the importance of robust image preprocessing.
9. **State-of-the-Art Comparison:** ResNet-18 surpassed several state-of-the-art models, including UNet+DenseNet201 and Vision Transformers, both in terms of accuracy and computational efficiency. Its simplicity allowed it to achieve high performance without the complexity of hybrid or ensemble models.

### 6.2) Future Work

1. **Real-time Clinical Integration:** Deploying these models into clinical environments could be further developed, integrating them with existing diagnostic systems to provide real-time decision support. This would aid radiologists in making faster and more accurate diagnoses, with fine-tuned models to improve precision in edge cases and reduce false positives/negatives.
2. **Cross-Modal Learning:** Future research could explore cross-modal learning by combining X-ray images with other types of medical data, such as patient demographic information or clinical reports. This multi-modal approach could enhance the model’s performance by providing additional context for better decision-making.

### 6.3) Conclusion

This study demonstrates that the ResNet-18 model, when applied to multiclass classification of chest X-ray images, outperforms DenseNet-121 and other state-of-the-art models in terms of accuracy, generalization, and computational efficiency. By incorporating a range of datasets, including Kermany, RSNA, and NIAID, and considering the inclusion of an “Non-X-ray” class to account for non-X-ray images, the model shows robust performance across diverse image types. The inclusion of class weights did not significantly improve performance, suggesting that the dataset’s inherent characteristics did not benefit substantially from such modifications.

ResNet-18, achieving a test accuracy of 98.76%, excelled in handling complex classes such as Pneumonia, Tuberculosis, and Non-X-ray, delivering high precision and recall scores. In comparison to any other study, which demonstrated a lower performance across key metrics, ResNet-18’s simpler architecture and computational efficiency make it particularly suitable for real-world deployment, especially in resource-constrained environments. Additionally, its adaptability to various data sources and imaging conditions further highlights its potential for use in clinical applications.

The findings of this study underscore the importance of selecting the right model for practical deployment, with ResNet-18 emerging as the preferred choice due to its balance of performance, efficiency, and adaptability. The insights from this research provide a foundation for future work, particularly in exploring the expansion of the “Non-X-ray” category to further improve model robustness, and the exploration of more advanced optimization techniques to enhance model generalization.

## Data Availability

All data used in this study were anonymized and publicly available for research purposes. No additional ethical approval was required as the datasets utilized are open-access, and their use for research has been authorized by the original data sources. The study was conducted in accordance with the Declaration of Helsinki.

https://www.kaggle.com/competitions/rsna-pneumonia-detection-challenge

https://www.kaggle.com/datasets/pavansanagapati/images-dataset/data

